# Impact of DENV-ZIKV cross-reactivity in Southeast Asia: A modelling study in Thailand

**DOI:** 10.64898/2026.11.08.26367564

**Authors:** Yile Shi, Abhishek Senapati, Angkana T. Huang, Hannah E. Clapham

**Affiliations:** Saw Swee Hock School of Public Health, National University of Singapore and National University Health System, Singapore

**Author notes:** Author for correspondence: Hannah E. Clapham.

**Keywords:** dengue virus, Zika virus, cross-reactivity, cross-protection, immune enhancement, mathematical modelling, population ageing, surveillance

## Abstract

Dengue virus (DENV) is endemic across Southeast Asia (SEA), where a large proportion of adults have been infected. Zika virus (ZIKV) has circulated in the region for decades but the extent of transmission remains unclear. Prior infection with one virus can protect against or enhance infection with the other, but the population-level consequences are unknown. We developed an age-structured DENV-ZIKV model with nine cross-reactivity assumptions combining cross-protection and subsequent enhancement, and simulated sustained co-circulation under changing Thai demography. Through cross-reactivity, DENV strongly shaped ZIKV transmission, whereas ZIKV had little effect on DENV transmission. Under cross-protection, ZIKV transmission occurred as infrequent outbreaks without changes in long-term ZIKV burden. When prior DENV infection enhanced both ZIKV infection and symptomatic risk, this burden roughly doubled. Population ageing raised the mean age of DENV and ZIKV cases by over 20 years, more than cross-reactivity did. Simulated ZIKV case ages were closer to those reported in Thailand in 2019–2023 under enhancement of ZIKV infection and symptoms. Over half of individuals aged 15–49 years remained ZIKV-naive under all assumptions, leaving many pregnancies at risk of congenital Zika syndrome. Interpreting ZIKV case counts in SEA requires joint consideration of cross-reactivity, demographic change and case detection.

## 1. Introduction

Dengue virus (DENV) and Zika virus (ZIKV) are closely related mosquito-borne flaviviruses transmitted primarily by *Aedes* mosquitoes [1,2]. Dengue has expanded geographically and increased in incidence over recent decades, with hundreds of millions of estimated infections each year [1,3]. DENV infection may be asymptomatic or present as an acute febrile illness [1]. ZIKV infection is also frequently asymptomatic or clinically mild. Its severe outcomes depend on age at infection, with Guillain-Barré syndrome in adults and congenital Zika syndrome, including microcephaly, after infection during pregnancy [4,5]. Large outbreaks in the Americas in 2015–2016 drew wider attention to these consequences [2].

Cross-reactivity is well established among DENV serotypes but remains unresolved between DENV and ZIKV. Infection with one DENV serotype generates long-lasting homotypic immunity [6]. Heterotypic immunity involves temporary cross-protection against the other serotypes, followed by antibody-dependent enhancement (ADE), which can increase the risk of subsequent infection or severe disease [6,7]. Shared conserved antigenic features in the envelope protein of DENV and ZIKV allow cross-reactive antibodies from DENV-exposed individuals to bind or neutralize ZIKV, or to enhance infection at sub-neutralizing concentrations in vitro [8,9]. Animal studies provided inconsistent evidence of DENV-ZIKV cross-reactivity. In mice, pre-existing anti-DENV antibodies increased ZIKV infection and ZIKV-associated pregnancy damage [10,11]. However, some macaque studies found no clear effect of pre-existing DENV immunity on ZIKV infection or disease severity [12,13]. In the other direction, prior ZIKV exposure was found to increase DENV viraemia or disease severity in mice and macaques [14,15].

Evidence in humans supporting DENV-ZIKV cross-reactivity exists but is mostly from Latin America during the 2015–2016 ZIKV outbreak. Individual-level cohort and community studies in Nicaragua used antibody measurements to determine each person’s history of DENV and ZIKV infection. High anti-DENV antibody titres were associated with protection against ZIKV infection and symptomatic disease [16,17]. Results from a paediatric cohort showed that prior ZIKV infection increased the subsequent risk of severe dengue hospitalization [18]. Population-level modelling work in Brazil and Colombia suggested that temporary cross-protection induced by prior ZIKV exposure can explain the unusually low dengue incidence after ZIKV emergence [19,20]. In Brazil, dengue cases also became younger after the ZIKV epidemic, particularly where ZIKV attack rates were higher [21]. These findings suggest that DENV-ZIKV cross-reactivity can affect epidemic timing, case numbers and the age of cases.

In contrast, the magnitude of ZIKV outbreaks in Southeast Asia (SEA) was far more limited than in the Americas. In 2016, an outbreak in Singapore involved 455 confirmed cases, and dedicated surveillance in southern Vietnam identified 214 ZIKV-positive cases [22,23]. The limited reports in SEA could reflect reduced clinical expression in a DENV-immune population, limited transmission of ZIKV, or incomplete detection [24–26]. DENV is endemic in many SEA countries, and most adults have been infected at least once [27]. As populations across the region age, DENV immunity is retained by a growing share of older adults [28,29]. ZIKV has been documented sporadically in the region for decades [24]. In Thailand, routine surveillance has reported ZIKV cases consistently in recent years, including 818 cases in 2023 [30]. Phylogenetic evidence indicates that ZIKV has circulated there over the long term [31]. Serological surveys in asymptomatic populations suggest that ZIKV exposure is more common than reported cases indicate [32]. These surveys are difficult to interpret, as antibodies from prior DENV infection can cross-react with ZIKV in serological assays, particularly after repeated DENV exposure [8,25]. The combined effects of DENV-ZIKV cross-reactivity and population ageing on transmission have not been examined in a DENV-endemic SEA setting.

To investigate how DENV-ZIKV cross-reactivity and demographic change shape transmission in SEA, we developed an age-structured model framework that tracks infection histories across four DENV serotypes and ZIKV, basing our simulations on DENV epidemiology and demography in Thailand. We simulated sustained DENV-ZIKV co-circulation consistent with evidence of long-term ZIKV circulation [31]. We explored how different combinations of cross-protection and enhancement shape the transmission, age distribution and accumulation of immunity for both viruses under ageing demography. Comparing simulated ZIKV case ages with those reported in Thailand, we examined the extent to which observed age patterns could distinguish between cross-reactivity assumptions, and what reported cases indicate about ZIKV transmission in the region. Finally, we discuss possible explanations for the contrast between the limited ZIKV activity reported in SEA and the large epidemics in the Americas.

## 2. Methods

### 2.1 Model framework and cross-reactivity assumptions

The model tracks infection and immunity to each DENV serotype and to ZIKV by age. DENV infection with one serotype generates lifelong homotypic immunity and temporary heterotypic cross-protection against the other serotypes [6]. Once this temporary protection wanes, the risk of subsequent heterotypic infection and symptomatic illness is modified by enhancement depending on the assumption [7]. Individuals are assumed completely immune to DENV after infections with two distinct serotypes. ZIKV is modelled as a virus distinct from DENV, not as a fifth DENV serotype. ZIKV infection generates lifelong immunity to ZIKV. Each individual can therefore have up to three infections, two with distinct DENV serotypes and one with ZIKV, in any order. Complete immunity to both viruses requires all three infections. Simultaneous infections with multiple DENV serotypes or with both viruses are not considered. Sequential infection pathways are shown in Figure 1a, with transition equations provided in Supplementary material 1, section 4.

**Figure 1.**
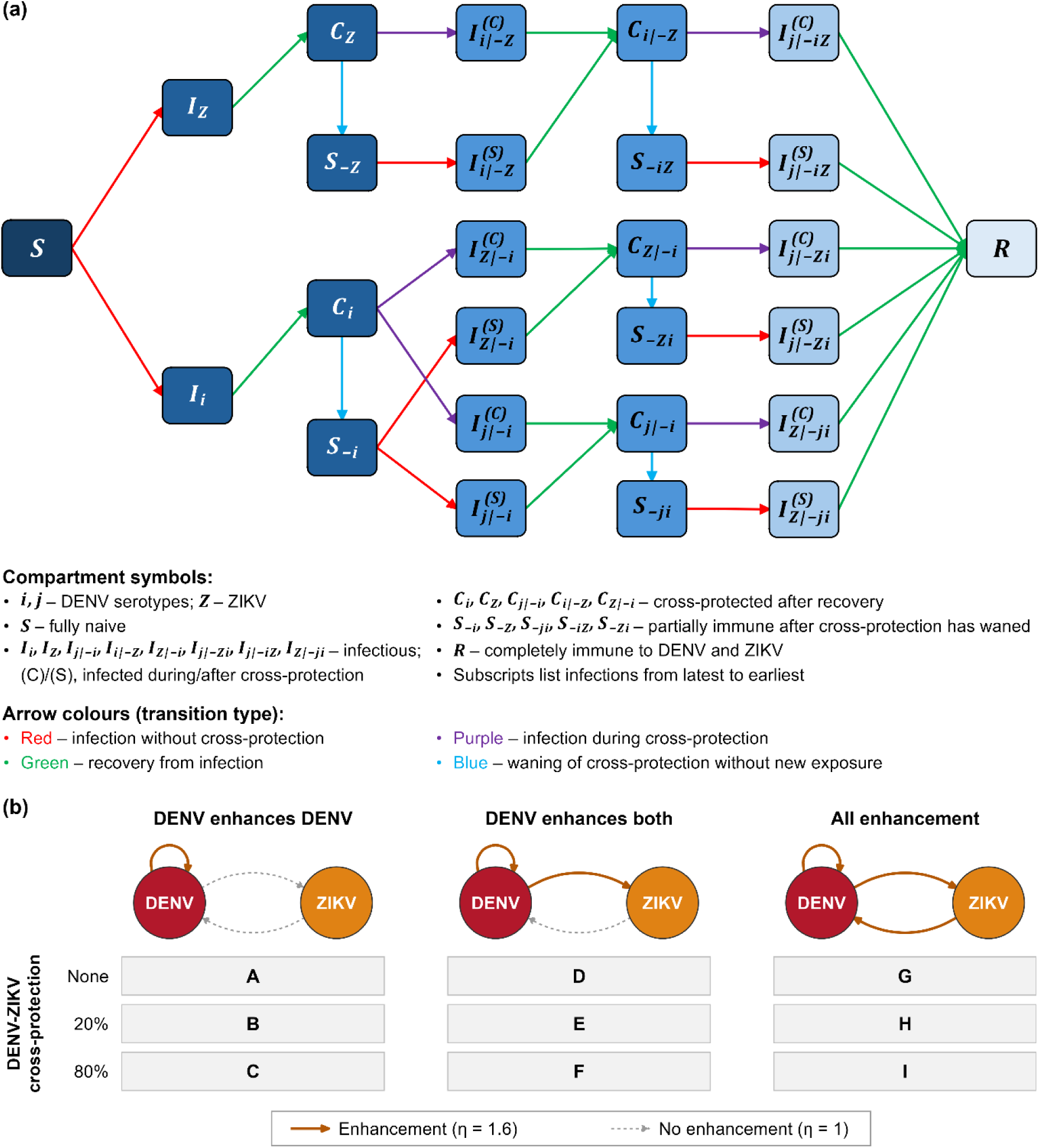
Model structure and DENV-ZIKV cross-reactivity assumptions. (a) Representative sequential infection pathways for DENV and ZIKV. Boxes denote compartments; subscripts list current and prior infections, and superscripts indicate infection during or after cross-protection. Arrow colours denote transition types. The pathways shown apply within each age group. Births, ageing and deaths are not shown. (b) The nine cross-reactivity assumptions (A-I). Columns show the three enhancement configurations, with enhanced directions drawn as solid arrows. Rows show no, weak (20%) and strong (80%) DENV-ZIKV cross-protection. The enhancement factor η is defined in Table 1.

**Table 1.** Model parameters and values used in the simulations. *Derived from the basic reproduction numbers, recovery rate and infectivity using an age-aggregated next-generation matrix approximation for primary infection under a fully susceptible state [36]. Infectivity rate is fixed at 1 d^−1^, so transmission intensity is captured by susceptibility (Supplementary material 1, section 5).

| Parameter | Description | Value | Reference |
|---|---|---|---|
| R<sub>0,D</sub> | Basic reproduction number for DENV | 3 | [27] |
| R<sub>0,Z</sub> | Basic reproduction number for ZIKV | 1.5 | [22,33] |
| σ | Recovery rate | 0.14 d<sup>−1</sup> | [1] |
| ϕ<sub>D</sub> | Infectivity rate of a DENV-infectious host | 1 d<sup>−1</sup> | Fixed |
| ϕ<sub>Z</sub> | Infectivity rate of a ZIKV-infectious host | 1 d<sup>−1</sup> | Fixed |
| ρ<sub>D</sub> | Baseline susceptibility to DENV | 0.43 | Computed* |
| ρ<sub>Z</sub> | Baseline susceptibility to ZIKV | 0.21 | Computed* |
| p<sub>D,pri</sub> | Symptomatic risk of primary DENV infection, without DENV-ZIKV cross-reactivity | 0.18 | [34] |
| p<sub>D,sec</sub> | Symptomatic risk of secondary DENV infection, without DENV-ZIKV cross-reactivity | 0.41 | [34] |
| p<sub>Z,cb</sub> | Symptomatic risk of ZIKV infection in pregnant women of childbearing age (15–49 years), without DENV-ZIKV cross-reactivity | 0.23 | [35] |
| p<sub>Z,g</sub> | Symptomatic risk of ZIKV infection in the general population, without DENV-ZIKV cross-reactivity | 0.18 | Assumed |
| w<sub>D</sub> | Waning rate of DENV-induced cross-protection | 1 y<sup>−1</sup> | [6] |
| w<sub>Z</sub> | Waning rate of ZIKV-induced cross-protection | 1 y<sup>−1</sup> | Assumed |
| κ<sub>DD</sub> | Susceptibility modification factor for heterotypic DENV infection during DENV-induced cross-protection, fixed at 80% protection | 0.2 | [19] |
| κ<sub>ZD</sub> | Susceptibility modification factor for DENV infection during ZIKV-induced cross-protection, explored at no, 20% and 80% protection (Figure 1b) | 1, 0.8, 0.2 | Explored |
| κ<sub>DZ</sub> | Susceptibility modification factor for ZIKV infection during DENV-induced cross-protection, explored at no, 20% and 80% protection (Figure 1b) | 1, 0.8, 0.2 | Explored |
| η<sub>ij</sub> | Susceptibility enhancement factor for heterotypic or cross-virus infection after cross-protection wanes | 1.6 | [19] |
| ξ<sub>ZD</sub> | Symptomatic risk modification factor for DENV infection after ZIKV infection, depending on the cross-reactivity assumption (section 2.2) | 1.6, 1 | Assumed |
| ξ<sub>DZ</sub> | Symptomatic risk modification factor for ZIKV infection after DENV infection, depending on the cross-reactivity assumption and symptomatic parameterization (section 2.2) | 1.6, 1; 0.62 | Assumed; [19] |

DENV-ZIKV cross-reactivity is represented by temporary cross-protection and enhancement. Cross-protection reduces susceptibility to subsequent heterotypic or cross-virus infection during the protected period, which lasts on average one year (Table 1). DENV-ZIKV cross-protection is assumed symmetric as a modelling simplification, and no, weak (20% reduction in susceptibility), and strong (80% reduction in susceptibility) protection levels are explored (Table 1). Enhancement increases susceptibility to subsequent heterotypic or cross-virus infection after the protection period wanes. We consider three enhancement configurations: enhancement of secondary heterotypic DENV infection only, additional enhancement of ZIKV infection after DENV infection, and further enhancement of DENV infection after ZIKV infection (Figure 1b). Combining the three enhancement configurations with the three DENV-ZIKV cross-protection levels gives nine DENV-ZIKV cross-reactivity assumptions, labelled A–I (Figure 1b).

We benchmark our model against Borchering et al. under comparable settings to check whether it captures DENV dynamics and the effect of DENV-ZIKV cross-reactivity on them [19]. The model captures the overall pattern of post-ZIKV DENV dynamics in their simulations (Figure S1).

### 2.2 Thai demographic setting and transmission parameterization

We use annual crude birth rates and age-specific mortality rates for Thailand from the World Population Prospects series for 1950–2100 to generate a time-varying population [28]. These comprise estimates for 1950–2023 and medium-variant projections for 2024–2100. The population is stratified into single-year age groups (0 to 89 years and 90+). Births enter the youngest age group as immunologically naive individuals, ignoring maternal immunity.

Transmission is represented among human hosts and vector dynamics are not modelled. All four DENV serotypes share the same transmission settings (Table 1). The basic reproduction number for DENV is set to 3 to represent endemic transmission intensity in Thailand [27]. In the absence of direct estimates for most SEA settings, the basic reproduction number for ZIKV in Thailand is set to 1.5, consistent with estimates from the 2016 Singapore outbreak (1.5–2) and from land-based populations in Latin America, such as Colombia and Guatemala (1.05– 1.75) [22,33]. DENV symptomatic probability differs between primary and secondary infections [34]. ZIKV symptomatic probability is specified separately for pregnant women aged 15–49 years, based on the 2016 French Guiana outbreak [35]. The proportion of pregnant women in each age group (15–49 years) is approximated from Thai age-specific fertility rates (Supplementary material 1, section 4.4) [28].

Simulated DENV and ZIKV cases are symptomatic infections, calculated from new infections according to infection history. Symptomatic risks change the number of cases and relevant age patterns but not infections or immunity. Between DENV serotypes, the effect of prior infection on symptomatic risk is included in the symptomatic risk of secondary DENV infection (p_D,sec_). Between DENV and ZIKV, the symptomatic risk of infection after infection with the other virus is that of primary infection multiplied by a factor ξ (Table 1). For DENV infection after ZIKV infection, ξ_ZD_ is assumed to equal the enhancement factor η under ZIKV-to-DENV enhancement (assumptions G–I) and 1 otherwise. For ZIKV infection after DENV infection, ξ_DZ_ is 1 without DENV-to-ZIKV enhancement (assumptions A–C). Under DENV-to-ZIKV enhancement (assumptions D–I), we consider two parameterizations of ξ_DZ_. In the enhanced symptomatic parameterization, ξ_DZ_ equals η. In the reduced symptomatic parameterization, ξ_DZ_ is 0.62, based on the lower risk of symptomatic ZIKV infection among children with prior DENV infection in a cohort in Nicaragua [17]. Results are reported for the enhanced symptomatic parameterization and for the reduced parameterization where the two differ.

### 2.3 Simulation design and outcome measures

We simulate sustained DENV-ZIKV co-circulation over 150 years (1950–2099) to cover the Thai demographic transition from a young to an aged population. Simulations start from a 300-year warm-up (Supplementary material 1, section 6). We run 100 simulations per cross-reactivity assumption, with susceptibility sampled from a distribution centred on the value in Table 1 for each virus (Supplementary material 1, section 6).

We select three 10-year windows (1970s, 2020s and 2050s) to represent young-age, ageing and aged demographic structures in Thailand (Figure S2). Long-term burden is summarized as the average annual number of DENV and ZIKV cases per 100,000 population over 1950–2099. The coefficient of variation (CV) of annual cases measures year-to-year variability in DENV and ZIKV transmission, within each window and over the full period. The recurrence intervals (RI) measure the frequency of elevated transmission, defined as the average time from the start of one period with monthly case number above the 75th percentile of the simulated time series to the start of the next. Within each window, we summarize age-specific immune profiles for DENV (naive, monotypically immune after one prior DENV infection, or completely immune after two), for ZIKV (naive or immune) and for both viruses jointly (naive to both viruses, immune to DENV only, immune to ZIKV only, or immune to both). From these profiles, we calculate the overall seroprevalence of each virus. Mean ages of DENV and ZIKV cases under each cross-reactivity assumption are also calculated within each demographic structure. Each outcome is calculated for each run and summarized as the mean and 95% quantile range across runs.

### 2.4 Comparison with Thai surveillance data

Simulated age distributions of ZIKV cases are compared with reported Thai ZIKV cases from 2019–2023 [30]. Simulated cases are pooled over the 10 years of the ageing structure, which overlaps the surveillance period and has a similar population age structure [28]. Reported cases are pooled across 2019–2023, and year-specific distributions are retained to show between-year variation. Simulated and reported data are expressed as relative age-group proportions across nine age groups (0–4, 5–9, 10–14, 15–24, 25–34, 35–44, 45–54, 55–64 and ≥65 years). Comparing age-group proportions removes the effect of any reporting fraction that is constant across ages. The primary measure is the total variation distance (TVD), which quantifies the overall difference between two distributions on a scale from 0 (identical) to 1 (no overlap). For each run, TVD is calculated as half the sum of absolute differences between simulated and pooled reported age-group proportions. The root mean squared error (RMSE) of these proportions is also calculated as a secondary measure. Cross-reactivity assumptions are compared by mean TVD and RMSE across the 100 runs. For assumptions D–I, the comparison is made under both the enhanced and the reduced symptomatic parameterizations (section 2.2).

## 3. Results

### 3.1 Cross-reactivity shapes ZIKV transmission but has little effect on DENV

Over the full simulation period, DENV-ZIKV cross-reactivity affected ZIKV transmission much more than DENV transmission (Figure 2, Table S1). When there was neither cross-protection nor enhancement between DENV and ZIKV (assumption A), ZIKV transmission persisted at a low level, with an average of 131.5 cases per 100,000 population per year and a coefficient of variation (CV) of annual ZIKV cases of 0.51 (Figure 2c, Table S1). DENV-ZIKV cross-protection turned this low-level transmission into recurrent ZIKV outbreaks separated by periods of very low transmission (Figure 2a). Stronger cross-protection made ZIKV transmission fluctuate more, with the CV of annual ZIKV cases rising to 1.13 under weak cross-protection (assumption B) and 2.81 under strong cross-protection (assumption C) (Figure 2c). ZIKV outbreaks also became less frequent but reached higher peaks, with the recurrence intervals (RI) between them lengthening from 9.9 years under weak to 16.4 years under strong cross-protection (Figure 2a,b). ZIKV cases were thus concentrated in these fewer outbreaks, and the average annual ZIKV burden stayed close to that without cross-protection, at 131.4 and 138.1 cases per 100,000 population (Table S1).

**Figure 2.**
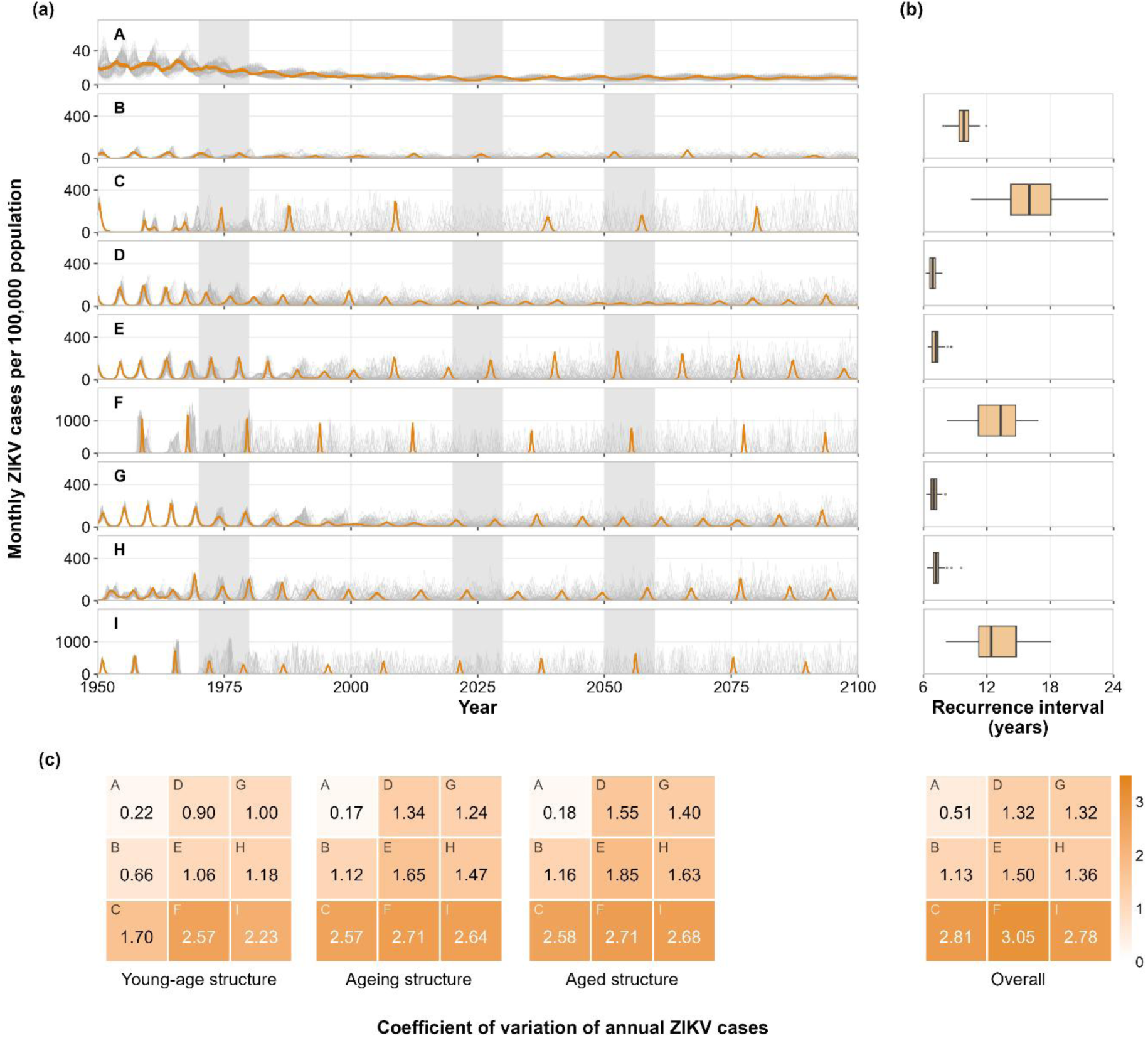
ZIKV transmission under sustained DENV-ZIKV co-circulation. Cases are shown for the enhanced symptomatic parameterization (section 2.2). (a) Monthly ZIKV cases per 100,000 population from 1950 to 2099 under each cross-reactivity assumption (A-I). Grey lines show individual runs and orange lines the run with the parameter values in Table 1. Shaded bands mark the three 10-year windows used for the young-age, ageing and aged demographic structures. (b) Box plots of the recurrence interval of ZIKV outbreaks across runs. Assumption A is not shown because it produced too few periods of elevated transmission to estimate a recurrence interval. (c) Heatmaps of the mean coefficient of variation of annual ZIKV cases across runs, within each demographic structure (below the corresponding windows in a) and over the full simulation period (Overall, below b). Heatmap cells are labelled by cross-reactivity assumption (Figure 1b).

When prior DENV infection also enhanced subsequent ZIKV infection (assumptions D–F), the average annual ZIKV burden in the enhanced symptomatic parameterization roughly doubled, to 270–285 cases per 100,000 population, relative to the burden without DENV-ZIKV cross-reactivity (Table S1). Under this enhancement, individuals with prior DENV infection were more susceptible to ZIKV and, in this parameterization, more likely to develop symptoms once infected. In the reduced symptomatic parameterization, the average annual ZIKV burden under assumptions D–F stayed close to that without DENV-ZIKV cross-reactivity, at 129–137 cases per 100,000 population (Figure S3, Table S1). In both symptomatic parameterizations, even without DENV-ZIKV cross-protection (assumption D), DENV-to-ZIKV enhancement turned ZIKV transmission into recurrent outbreaks, with a CV of annual ZIKV cases of 1.32 and an RI of 6.9 years (Figures 2 and S3). Weak cross-protection changed this pattern little (assumption E), but strong cross-protection again made ZIKV outbreaks less frequent, raising the CV of annual ZIKV cases to 3.05 and the RI to 13.1 years (assumption F) (Figure 2). When prior ZIKV infection also enhanced subsequent DENV infection (assumptions G–I), ZIKV transmission was very similar to that with DENV-to-ZIKV enhancement alone (Figure 2, Table S1). This additional enhancement acted only on subsequent DENV infection, so it could affect ZIKV transmission only indirectly, through DENV transmission.

By contrast, DENV transmission itself changed little across DENV-ZIKV cross-reactivity assumptions (Figure S4). The average annual DENV burden stayed at about 780–820 cases per 100,000 population (Table S1). When prior ZIKV infection raised both the susceptibility to DENV and the probability of symptomatic DENV infection (assumptions G–I), the DENV burden was about 3% higher than under assumptions A–F (Table S1). DENV epidemics recurred about every 4 years, with a CV of annual DENV cases of 1.2–1.3 (Figure S4). In all three demographic structures, over 80% of the population had at least one prior DENV infection (Figures S5–S7). Prior ZIKV infection was less common, at about 30% of the population without DENV-to-ZIKV enhancement and about 50% with it (Figures S8–S10). DENV-ZIKV cross-reactivity modified susceptibility to one virus only in individuals previously infected with the other. With this asymmetric immune background, DENV-ZIKV cross-reactivity acted on ZIKV infection in most of the population, but on DENV infection only in a smaller share.

### 3.2 Population ageing shifts the age of DENV and ZIKV cases more than cross-reactivity does

Population ageing shifted immunity and infection towards older ages for both viruses, regardless of DENV-ZIKV cross-reactivity assumption (Figures 3 and S11–S13). From the young-age to the ageing and aged structures, the mean age of the population rose from 23 to 41 and 50 years (Figure S2). The mean age of DENV cases rose accordingly, from about 9 years in the young-age structure to 22 and 30 years in the ageing and aged structures, and that of ZIKV cases in the enhanced symptomatic parameterization from 15–19 to 32–35 and 42–45 years (Figure 4, Table S2). In each demographic structure, the mean age of ZIKV cases was about 4–9 years below that of the whole population, whereas that of DENV cases was 14–20 years below it (Figure 4, Table S2). ZIKV immunity was acquired later as the population aged, and the proportion of ZIKV-naive individuals aged 15–49 years rose from about 30–60% in the young-age structure to 55–75% in the ageing structure (Figures S8–S10).

**Figure 3.**
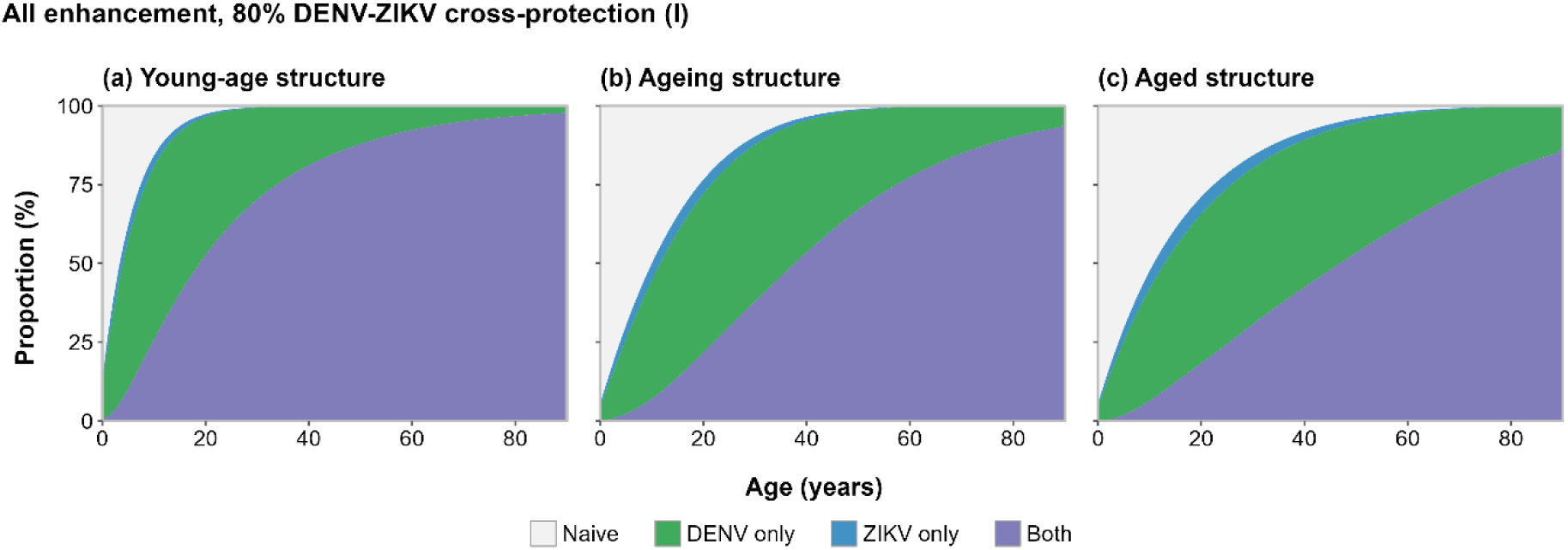
Age-specific immunity to DENV and ZIKV across demographic structures under sustained co-circulation. Panels show the proportions of individuals who were naive (grey), immune to DENV only (green), immune to ZIKV only (blue) or immune to both viruses (purple) by age, under all enhancement with strong DENV-ZIKV cross-protection (assumption I). Panels (a), (b) and (c) show the young-age, ageing and aged structures, based on Thai demographic parameters for the 1970s, 2020s and 2050s. Proportions are means across 100 runs.

**Figure 4.**
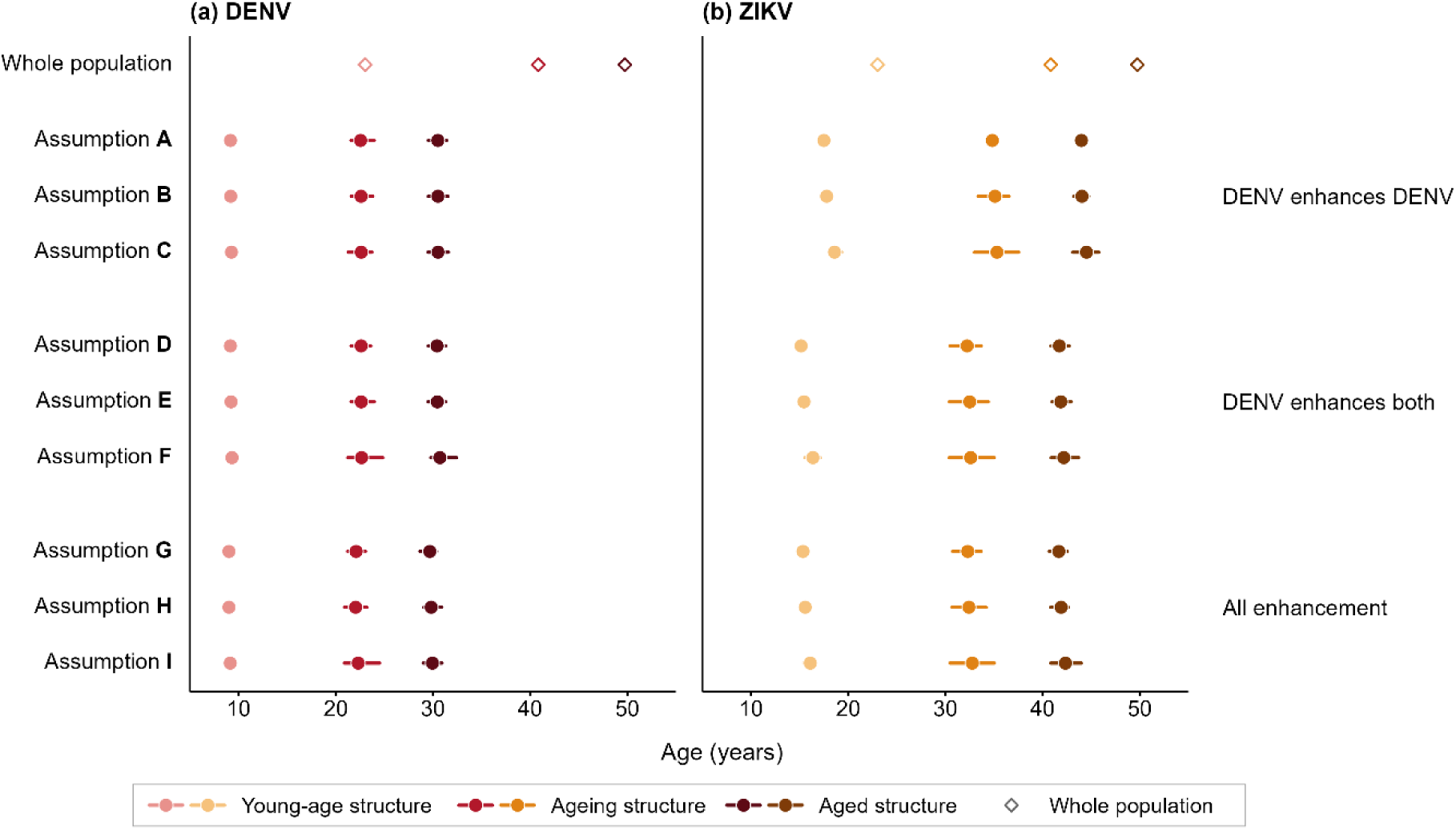
Mean age of DENV and ZIKV cases under co-circulation. ZIKV cases are shown for the enhanced symptomatic parameterization (section 2.2). Points show the mean age of (a) DENV and (b) ZIKV cases across runs under each cross-reactivity assumption, and horizontal lines show the 95% quantile range. Colour shades denote the young-age, ageing and aged demographic structures, based on Thai demographic parameters for the 1970s, 2020s and 2050s. Open diamonds show the mean age of the whole population in each structure. Assumptions are grouped by enhancement configuration (Figure 1b). Values are given in Table S2.

Population ageing changed these age patterns far more than DENV-ZIKV cross-reactivity did (Figure 4). Within each demographic structure, the mean age of DENV cases differed by about 1 year or less across DENV-ZIKV cross-reactivity assumptions and that of ZIKV cases by less than 3.5 years in the enhanced symptomatic parameterization, compared with increases of about 21 and 26 years from the young-age to the aged structure. The differences in ZIKV case age came mainly from DENV-to-ZIKV enhancement, under which ZIKV immunity was acquired earlier and ZIKV cases were 2–3 years younger in each demographic structure (Figures S11–S13). In the reduced symptomatic parameterization, ZIKV cases under assumptions D–I were a further 2–4 years younger (Figure S14). In this parameterization, the mean age of ZIKV cases differed by up to about 7 years across assumptions within each demographic structure, which remained far smaller than the rise of over 20 years with population ageing (Figure S14). Population ageing shifted the age distribution of individuals susceptible to each virus, whereas DENV-ZIKV cross-reactivity changed their susceptibility within this distribution.

Besides shifting infection towards older ages, population ageing made transmission more variable from year to year. For DENV, the CV of annual cases rose from about 0.8–1.1 in the young-age structure to 1.4–1.7 in the ageing and aged structures under all assumptions (Figure S4). This increase was larger than the differences in the CV of annual DENV cases across cross-reactivity assumptions within each structure. For ZIKV, the corresponding CV rose by about 0.1–0.9 from the young-age to the aged structure under DENV-ZIKV cross-reactivity (assumptions B–I) (Figure 2c). Without this cross-reactivity (assumption A), the CV of annual ZIKV cases stayed at about 0.2 in all three structures (Figure 2c).

### 3.3 Thai ZIKV case ages are closer to simulations in which prior DENV infection enhances ZIKV infection and symptoms

In the enhanced symptomatic parameterization, simulated age distributions of ZIKV cases were closer to the reported Thai distribution when prior DENV infection enhanced subsequent ZIKV infection (assumptions D–I) than without this enhancement (assumptions A–C) (Figure 5, Table S3). The TVD from the pooled reported distribution was 0.146–0.150 with DENV-to-ZIKV enhancement and 0.159–0.161 without it, and the RMSE ranked the DENV-ZIKV cross-reactivity assumptions in the same order (Table S3). Simulated age distributions largely overlapped among assumptions with DENV-to-ZIKV enhancement (assumptions D–I), and among those without it (assumptions A–C) (Figure 5). TVD differed by less than 0.005 among assumptions with DENV-to-ZIKV enhancement, which is consistent with the small effects of cross-protection and ZIKV-to-DENV enhancement on the mean age of ZIKV cases (Figure 4, Table S3).

**Figure 5.**
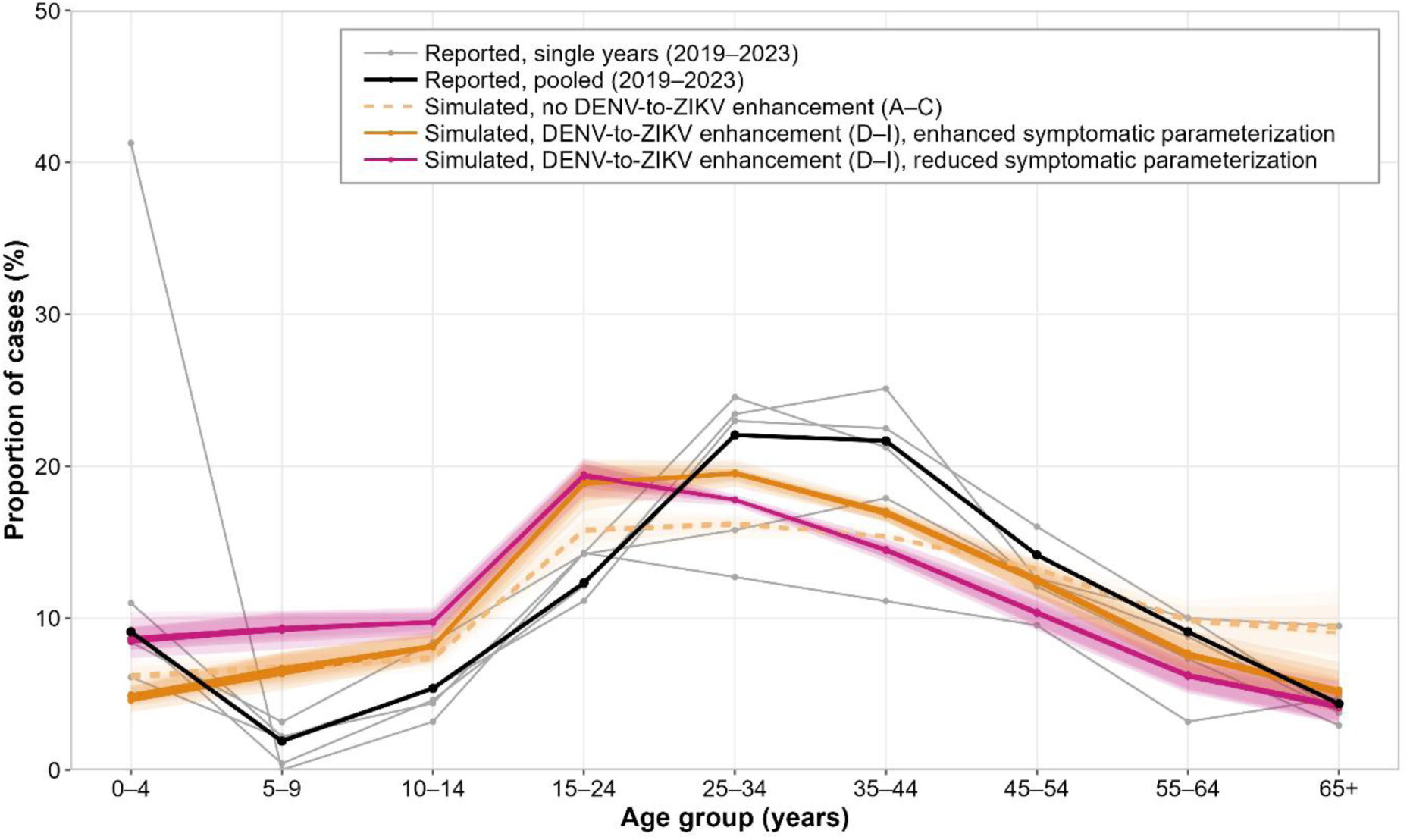
Simulated and reported age distributions of ZIKV cases in Thailand. Grey lines show the age distributions of reported ZIKV cases in each year from 2019 to 2023. The black line shows the age distribution pooled across the five years. Coloured lines show the mean age distribution of simulated ZIKV cases in the ageing structure across 100 runs, with one line for each cross-reactivity assumption (Figure 1b). Dashed lines show the three assumptions without DENV-to-ZIKV enhancement (assumptions A–C), which are identical in the two symptomatic parameterizations. Solid lines show the six assumptions with this enhancement (assumptions D–I) in the enhanced (orange) and reduced (magenta) symptomatic parameterizations (section 2.2). Lines of the same colour largely overlap. Shading shows the 95% quantile range across runs for each assumption. Reported data are from [30] and provided in Table S4.

Cases aged 15–44 years made up 56% of reported cases, compared with about 55% of simulated cases with DENV-to-ZIKV enhancement and about 47% without it (Figures 5 and S15). Within this age range, simulated cases with DENV-to-ZIKV enhancement were concentrated at younger ages than reported cases, with a higher share at 15–24 years and a lower share at 35–44 years (Figure 5). Simulated distributions under all assumptions were also lower at 0–4 years, an age group in which infants under 1 year alone made up 7% of all reported cases (Figure 5, Table S4). The age distribution of reported cases varied between years, with the share of cases aged 15–44 years ranging from 48% to 61% in all years except 2021 (Figure 5, Table S4). In 2021, infants made up 40% of the 63 cases reported that year (Table S4).

In the reduced symptomatic parameterization, simulated ZIKV case ages under assumptions D–I were further from the reported distribution than those under assumptions A–C (Figure 5, Table S3). The TVD was 0.189–0.193 under assumptions D–I, compared with 0.159–0.161 under assumptions A–C, and the RMSE ranked the assumptions in the same order (Table S3).

Simulated cases under assumptions D–I were younger in this parameterization, with a higher share at 0–4 years and a lower share above 25 years than in the enhanced symptomatic parameterization (Figures 5 and S15). In this parameterization, ZIKV infections in individuals with prior DENV infection, most of whom were adults, were less likely to be symptomatic than those in DENV-naive individuals, most of whom were children (Figure 3).

## 4. Discussion

In DENV-endemic Southeast Asia, both the extent of ZIKV transmission and the population-level effects of DENV-ZIKV cross-reactivity on either virus remain uncertain. Our framework, which models the two-way cross-reactivity between DENV and ZIKV and extends previous DENV-ZIKV models, projected that cross-reactivity mainly affected ZIKV transmission and changed DENV transmission little [19,21]. In our model, population ageing changed the age of DENV and ZIKV cases more than DENV-ZIKV cross-reactivity did. The age distribution of reported ZIKV cases in Thailand was more consistent with model results in which prior DENV infection enhanced ZIKV infection and raised the risk of symptoms.

In settings with high DENV transmission, four DENV serotypes circulate against a single ZIKV serotype, so DENV immunity builds up across repeated infections and reaches most of the population, whereas ZIKV immunity remains limited [27,32]. In our model, DENV-ZIKV cross-protection changed the timing of ZIKV transmission, turning it into infrequent outbreaks separated by long periods of very low transmission (assumptions B, C, E, F, H and I), but kept long-term ZIKV burden close to that without cross-protection (assumptions A, D and G). Between DENV serotypes, temporary cross-protection is known to generate multiannual DENV epidemic cycles [37]. Our results suggest that temporary DENV-ZIKV cross-protection has a similar effect on ZIKV transmission. A ZIKV outbreak after many years of low activity could arise from sustained circulation, as in our model, and does not by itself indicate a new introduction. Genomic data would be needed to distinguish such recurrences from new introductions. The long-term ZIKV burden roughly doubled only when prior DENV infection enhanced both ZIKV infection and its symptoms (assumptions D–I in the enhanced symptomatic parameterization). However, across all assumptions, our model projected that long-term DENV burden stayed within about 3% of that without DENV-ZIKV cross-reactivity (assumption A). This suggests that the presence of ZIKV is unlikely to affect endemic dengue over the long term. Earlier modelling linked post-ZIKV dengue declines in Brazil and Colombia to temporary cross-protection from ZIKV infection [19,20]. Those declines followed outbreaks that infected a large share of the population within a short period, so many individuals were cross-protected against DENV at the same time [38]. In our model, under sustained co-circulation, ZIKV infections were spread over time, so fewer individuals were cross-protected against DENV at any one time. Where ZIKV circulates continuously, as phylogenetic evidence suggests for Thailand, dengue trends may show little signal of ZIKV transmission [31].

DENV-ZIKV cross-reactivity acts through prior immunity to the other virus, and population ageing shifted the age at which the immunity was acquired. In Thailand, the mean age of reported dengue haemorrhagic fever cases rose from 8.1 years in 1981 to 24.3 years in 2017, and a modelling study estimated that shifts in the age structure of susceptibility explained 58% of this rise [29]. In our simulations, with transmission parameters fixed, the mean age of DENV cases rose by a comparable amount, from about 9 years in the young-age structure to 22 years in the ageing structure. A rise of this size in the age of dengue cases is possible without any decline in transmission. Ageing shifted susceptibility to older ages and raised the age of cases for both viruses. DENV-ZIKV cross-reactivity changed only the susceptibility of individuals with a given infection history within the age structure shaped by ageing, shifting the age of DENV and ZIKV cases by up to about 7 years across assumptions A–I and both symptomatic parameterizations. Ageing also slowed the supply of new DENV-susceptible individuals through births, so DENV transmission relied more on periodic epidemics and became more variable over time. This is consistent with the longer intervals between dengue epidemics associated with declining birth rates in Thailand [39]. Ageing had a similar effect on ZIKV transmission wherever DENV-ZIKV cross-reactivity produced recurrent outbreaks (assumptions B–I). Similar changes in age patterns and temporal variability may occur elsewhere in SEA, where populations are also ageing [28].

Projected ZIKV seroprevalence in our model was lower than DENV seroprevalence under all cross-reactivity assumptions (about 30–50% vs over 80%; Figures S5–S10). The mean age of ZIKV cases stayed closer to that of the whole population than the mean age of DENV cases did (Figure 4). As the population aged, ZIKV infection moved further into adulthood, and a growing share of women reached reproductive age without prior ZIKV infection. Even when prior DENV infection enhanced ZIKV infection (assumptions D–I), our model projected that over half of individuals aged 15–49 years remained ZIKV-naive in the ageing structure (Figures S8–S10). Most of these individuals had prior DENV infection (Figures 3b and S12). Under DENV-ZIKV cross-protection (assumptions B, C, E, F, H and I), these susceptible adults could accumulate during the long periods between ZIKV outbreaks. A subsequent outbreak could then infect many women of reproductive age within a short period. ZIKV-naive women may remain at risk of ZIKV infection during pregnancy, which can cause congenital Zika syndrome [5]. Further research is needed to determine the impact of prior dengue immunity specifically on the risk of congenital Zika syndrome. The shift of ZIKV infection towards adults, most of whom carry DENV immunity, also matters beyond pregnancy, as ZIKV infection in adults has been linked to Guillain-Barré syndrome [4].

The comparison of our model results with Thai data was informative about whether prior DENV infection enhanced ZIKV infection and symptoms. In vitro and mouse studies have shown that antibodies from prior DENV infection can enhance ZIKV infection, although studies in macaques found no such effect [9,10,12,13]. Our comparison is consistent with this mechanism that prior DENV infection can enhance subsequent ZIKV infection at the population level in DENV-endemic settings. Reported ZIKV case ages were closer to simulations with this enhancement only when it also raised the risk of symptoms. With the lower symptomatic risk after DENV infection, simulated ZIKV cases were younger and further from the reported ages than without enhancement. DENV-ZIKV cross-protection and ZIKV-to-DENV enhancement had little effect on the age of ZIKV cases, so the comparison could not further identify which assumptions with these mechanisms best matched the reported data. Within ages 15–44 years, reported cases were older than simulated cases in both symptomatic parameterizations. This may reflect a higher symptomatic risk in adults with prior DENV infection than assumed, or age-dependent detection, such as differences in health-care seeking by age. Current surveillance data cannot distinguish between these explanations. Estimates of how DENV immune history affects symptomatic ZIKV infection in adults, who made up most reported cases in Thailand, would help interpret ZIKV case ages in SEA. The excess of reported cases among infants under 1 year may reflect the follow-up of ZIKV-infected pregnant women and their newborns in Thai surveillance [40,41]. The high infant share in 2021, a year with few reported cases, is consistent with this follow-up (Table S4). The age distribution of reported ZIKV cases in Thailand may therefore partly reflect who is tested. Surveillance of pregnant women and newborns remains essential for detecting congenital outcomes, but it cannot measure the extent of ZIKV infection in the population. Enhancement of ZIKV infection by maternal DENV antibodies could also contribute to the infant excess. Maternal immunity was not included in our model and could be assessed in future work.

DENV-to-ZIKV enhancement raised the proportion of the population with prior ZIKV infection from about 30% to about 50% (Figures S8–S10). These additional infections occurred mainly in individuals with prior DENV infection, and ZIKV immunity was acquired at younger ages (Figures S11–S13). If prior DENV infection lowers the risk of ZIKV symptoms, most of these infections would not be detected as cases [17]. In our reduced symptomatic parameterization, the long-term number of ZIKV cases stayed close to that without this enhancement (Table S1). ZIKV immunity could then build up earlier in life without an increase in reported cases. This is one plausible reason why no reported ZIKV circulation in DENV-endemic SEA has approached the scale of outbreaks in the Americas, where ZIKV spread through largely naive populations [38]. In other SEA settings, as in Singapore, sporadic infections and small clusters have been detected since the 2016 outbreak, but surveillance is limited by diagnostic capacity and the misclassification of ZIKV infections as dengue [24,42]. Serology does not resolve this in DENV-immune populations, as antibodies from prior DENV infection cross-react with ZIKV [24,25]. In our model results, the age of cases was shaped mainly by population structure, so comparisons of case age over time would require age standardization. Recording case age in single years would help detect the smaller shifts associated with DENV-to-ZIKV enhancement. Serological surveys that can separate ZIKV from DENV immunity would show how ZIKV immunity is distributed by age, including among women of reproductive age. Where ZIKV has not circulated continuously, new introductions into SEA populations with high DENV immunity remain possible. Future work could assess how DENV-ZIKV cross-reactivity and population ageing shape the consequences of such introductions.

The effect of prior ZIKV infection on dengue vaccine efficacy has yet to be established and should be considered [43]. In the phase 3 trial of the dengue vaccine TAK-003, children were enrolled in eight countries from September 2016 [44]. In the first year after vaccination, the placebo groups at most Latin American sites had few dengue cases, which increased in the following two years [44]. This period followed the 2015–2016 ZIKV epidemic, after which dengue incidence in the region was unusually low [19,20]. Temporary cross-protection from prior ZIKV infection may have contributed to the low case numbers, but this was not assessed in the trial. The effect of prior ZIKV infection on dengue vaccine efficacy may differ between settings and remains to be explored. In Thailand, where TAK-003 has been licensed, dengue vaccination would add vaccine-induced DENV immunity in a population with ongoing DENV and ZIKV transmission [45]. Whether vaccine-induced DENV immunity affects subsequent ZIKV infection, as infection-induced DENV immunity may do, is unknown. Data on both viruses from dengue vaccine studies would allow future work to examine these impacts across SEA within the same framework.

This work used a deterministic framework without explicit vector dynamics. Transmission parameters were set from previous estimates, and the strength and duration of cross-protection and enhancement were set from previous studies or varied across plausible levels to explore their population-level effects. Our results therefore show broad patterns in these effects. Future cohort estimates of the strength and duration of cross-protection and enhancement could be used within the same framework to quantify these effects.

Our findings provide model-based evidence on the population-level impacts of DENV-ZIKV cross-reactivity in a setting with high DENV transmission. In such a setting, DENV-ZIKV cross-reactivity can change the timing and size of ZIKV transmission while having little effect on dengue. In ageing populations, a growing share of adults may remain susceptible to ZIKV. In Thailand and other SEA settings with high DENV immunity and ageing populations, interpreting DENV and ZIKV case counts requires joint consideration of cross-reactivity, demographic change and case detection.

## Supporting information

Supplementary material 1: Model equations and derivations

## Data Availability

The model code, simulation summary outputs and the reported Thai ZIKV case data used in this study are available at https://github.com/Shi-Yile/denv-zikv-cross-reactivity-thailand (release v1.0). Demographic data are from the United Nations World Population Prospects 2024. Reported ZIKV cases are from the Department of Disease Control, Ministry of Public Health, Thailand.

https://population.un.org/wpp/

http://doe1.moph.go.th/surdata/disease.php?dcontent=old&ds=87

## Data accessibility

The model code, simulation summary outputs and reported Thai ZIKV case data used in this study are available at https://github.com/Shi-Yile/denv-zikv-cross-reactivity-thailand (release v1.0). Demographic data are from the United Nations World Population Prospects 2024 [28]. Reported ZIKV cases are from the Department of Disease Control, Ministry of Public Health, Thailand [30].

## Declare of AI use

Claude (Anthropic) was used to assist with literature searching, code debugging and language editing of the manuscript. The authors reviewed and verified all AI-assisted outputs and take full responsibility for the content of this article.

