## Supplementary material 1: Model equations and derivations for "Impact of DENV-ZIKV cross-reactivity in Southeast Asia: A modelling study in Thailand"

### 1. Model outline

The deterministic age-structured model describes the joint transmission dynamics of four DENV serotypes (DENV1-4) and one ZIKV serotype in a population structured by age and infection history (main text Figure 1a). Individuals are tracked through susceptible, infectious, temporary cross-protected, post-protection susceptible, and fully immune states. Infection history is tracked sequentially, such that indices $i$, $j$, and $k$ denote the infections in chronological order when they appear jointly. Demographic processes including birth, ageing, and natural mortality link successive age groups through inflows and outflows.

### 2. State variables and compartmental definitions

The model tracks individuals according to their infection and immunity history with respect to four DENV serotypes and ZIKV. State variables corresponding to the compartments in main text Figure 1a are defined below (Table A1).

All state variables are indexed by age group $a$, which is omitted from the definitions above for clarity. Superscripts $(C)$ and $(S)$ indicate infections occurring during and after the relevant cross-protection period, respectively. Each index is drawn from $\{1, 2, 3, 4, Z\}$. Tertiary infection is possible only when exactly one of $i$, $j$ and $k$ is ZIKV.

Numerical values for all parameters and details of cross-reactivity configurations are provided in main text Table 1 and Figure 1b.

| Symbol | Definition |
|---|---|
| $S^{[a]}$ | Fully susceptible individuals without prior infection by any DENV serotype or ZIKV. |
| $I_i^{[a]}$ | Individuals infected with virus or serotype $i$ as primary infection. |
| $C_i^{[a]}$ | Individuals in the temporary cross-protection period following primary infection with $i$. |
| $S_{-i}^{[a]}$ | Individuals after waning of cross-protection induced by primary infection with $i$, remaining susceptible to other virus or serotypes not yet experienced. |
| $I_{j\|-i}^{(C)[a]}$ | Individuals infected with $j$ during the cross-protection period after prior infection with $i$. |
| $I_{j\|-i}^{(S)[a]}$ | Individuals infected with $j$ after waning of cross-protection following prior infection with $i$. |
| $C_{j\|-i}^{[a]}$ | Individuals in the temporary cross-protection period after sequential infections with $i$ and then $j$. |
| $S_{-ji}^{[a]}$ | Individuals after waning of cross-protection following sequential infections with $i$ and then $j$, remaining susceptible to other virus or serotypes not yet experienced. |
| $I_{k\|-ji}^{(C)[a]}$ | Individuals infected with $k$ during the cross-protection period after prior sequential infections with $i$ and then $j$. |
| $I_{k\|-ji}^{(S)[a]}$ | Individuals infected with $k$ after waning of cross-protection following prior sequential infections with $i$ and then $j$. |
| $R^{[a]}$ | Individuals completely immune to DENV and ZIKV. |

Table A1: State variables and compartment definitions

### 3. Model parameters

The model parameters governing transmission, recovery, cross-reactivity, and demographic processes are described as follows in Table A2.

In $\kappa_{ij}$, $\eta_{ij}$ and $\xi_{ij}$, the first subscript denotes a prior infection and the second the current infection. Cross-protection depends on the most recent prior infection only. Enhancement from an earlier infection is retained after a later infection. After two prior infections, the larger of their enhancement factors applies once cross-protection has waned. In main text Table 1, the subscript $D$ denotes any DENV serotype, as all four serotypes share the same parameter values.

| Parameter | Definition |
|---|---|
| $\rho_i$ | Baseline susceptibility to infection with virus or serotype $i$. |
| $\phi_i$ | Infectivity rate of individuals currently infectious with virus or serotype $i$. |
| $\sigma$ | Recovery rate from infection. |
| $w_i$ | Waning rate of temporary cross-protection induced by infection with virus or serotype $i$. |
| $\kappa_{ij}$ | Susceptibility modification factor applied to baseline susceptibility to infection with virus or serotype $j$ during the temporary cross-protection period following prior infection with virus or serotype $i$. |
| $\eta_{ij}$ | Susceptibility modification factor applied to baseline susceptibility to infection with virus or serotype $j$ after the temporary cross-protection period following prior infection with virus or serotype $i$ has waned. |
| $\xi_{ij}$ | Symptomatic risk modification factor applied to the symptomatic probability of infection with virus or serotype $j$ after prior infection with virus or serotype $i$. Defined between DENV and ZIKV only (section 4.4). |
| $\mu$ | Per capita birth rate. |
| $m^{[a]}$ | Age-specific mortality rate. |
| $\alpha$ | Ageing rate linking successive age groups. |
| $N$ | Total population size. |

Table A2: Parameter definitions used in the transmission model

### 4. Model equations

For a given virus or serotype $v \in \{1, 2, 3, 4, Z\}$, the **force of infection (FOI)** is defined as

$$\lambda_v = \frac{\rho_v \phi_v}{N} \left[ \sum_a I_v^{[a]} + \sum_a \sum_{i \neq v} \left( I_{v|-i}^{(C)[a]} + I_{v|-i}^{(S)[a]} \right) + \sum_a \sum_{i \neq v} \sum_{\substack{j \neq v \\ j \neq i}} \left( I_{v|-ji}^{(C)[a]} + I_{v|-ji}^{(S)[a]} \right) \right]$$

State variables and parameters follow the definitions given above. All infectious individuals carrying virus or serotype $v$ contribute equally to the force of infection regardless of prior infection history, which means the infectivity parameter, $\phi_v$, depends on the current infecting virus or serotype and not on the order of previous infections. This formulation assumes homogeneous mixing across age groups and infection histories, such that all infectious individuals contribute proportionally to transmission according to their infectious status.

Using this force of infection, the compartmental dynamics in age group $a$ are given by the following system of ordinary differential equations (ODE).

#### 4.1 Primary infection and post-primary states

- **Fully susceptible individuals**

Transitions: $S^{[a]} \to I_i^{[a]}, \forall i$

$$\frac{dS^{[a]}}{dt} = -\sum_i \lambda_i S^{[a]} + \mathbb{1}_{a=0} \cdot \mu N + \mathbb{1}_{a\geq 1} \cdot \alpha S^{[a-1]} - \big(\mathbb{1}_{a<A} \cdot \alpha + m^{[a]}\big) S^{[a]}$$

- **Primary infection with virus or serotype $i$**

Transition: $I_i^{[a]} \to C_i^{[a]}$

$$\frac{dI_i^{[a]}}{dt} = \lambda_i S^{[a]} - \sigma I_i^{[a]} + \mathbb{1}_{a\geq 1} \cdot \alpha I_i^{[a-1]} - \big(\mathbb{1}_{a<A} \cdot \alpha + m^{[a]}\big) I_i^{[a]}$$

- **Temporary cross-protection following primary infection with $i$**

Transitions: $C_i^{[a]} \to I_{j|-i}^{(C)[a]} / S_{-i}^{[a]}$

$$\frac{dC_i^{[a]}}{dt} = -\sum_{j\neq i} \kappa_{ij} \lambda_j C_i^{[a]} - w_i C_i^{[a]} + \sigma I_i^{[a]} + \mathbb{1}_{a\geq 1} \cdot \alpha C_i^{[a-1]} - \big(\mathbb{1}_{a<A} \cdot \alpha + m^{[a]}\big) C_i^{[a]}$$

- **Post-protection susceptible state following primary infection with $i$**

Transition: $S_{-i}^{[a]} \to I_{j|-i}^{(S)[a]}$

$$\frac{dS_{-i}^{[a]}}{dt} = -\sum_{j\neq i} \eta_{ij} \lambda_j S_{-i}^{[a]} + w_i C_i^{[a]} + \mathbb{1}_{a\geq 1} \cdot \alpha S_{-i}^{[a-1]} - \big(\mathbb{1}_{a<A} \cdot \alpha + m^{[a]}\big) S_{-i}^{[a]}$$

#### 4.2 Secondary infection and post-secondary states

- **Secondary infection during cross-protection**

Transition: $I_{j|-i}^{(C)[a]} \to C_{j|-i}^{[a]}$

$$\frac{dI_{j|-i}^{(C)[a]}}{dt} = \kappa_{ij} \lambda_j C_i^{[a]} - \sigma I_{j|-i}^{(C)[a]} + \mathbb{1}_{a\geq 1} \cdot \alpha I_{j|-i}^{(C)[a-1]} - \big(\mathbb{1}_{a<A} \cdot \alpha + m^{[a]}\big) I_{j|-i}^{(C)[a]}$$

- **Secondary infection after waning of cross-protection**

Transition: $I_{j|-i}^{(S)[a]} \to C_{j|-i}^{[a]}$

$$\frac{dI_{j|-i}^{(S)[a]}}{dt} = \eta_{ij} \lambda_j S_{-i}^{[a]} - \sigma I_{j|-i}^{(S)[a]} + \mathbb{1}_{a\geq 1} \cdot \alpha I_{j|-i}^{(S)[a-1]} - \big(\mathbb{1}_{a<A} \cdot \alpha + m^{[a]}\big) I_{j|-i}^{(S)[a]}$$

- **Temporary cross-protection following secondary infection $j$**

Transition: $C^{[a]}_{j|-i} \to I^{(C)[a]}_{k|-ji} / S^{[a]}_{-ji}$ :

$$\frac{dC^{[a]}_{j|-i}}{dt} = -\sum_{\substack{k\neq i\\k\neq j}} \kappa_{jk}\eta_{ik}\lambda_k C^{[a]}_{j|-i} - w_j C^{[a]}_{j|-i} + \sigma\big(I^{(C)[a]}_{j|-i} + I^{(S)[a]}_{j|-i}\big) + \mathbb{1}_{a\geq 1}\cdot \alpha C^{[a-1]}_{j|-i} - \big(\mathbb{1}_{a<A}\cdot\alpha + m^{[a]}\big) C^{[a]}_{j|-i}$$

- **Post-protection susceptible state following secondary infection with $j$**

Transition: $S^{[a]}_{-ji} \to I^{(S)[a]}_{k|-ji}$

$$\frac{dS^{[a]}_{-ji}}{dt} = -\sum_{\substack{k\neq i\\k\neq j}} \max(\eta_{ik},\eta_{jk})\lambda_k S^{[a]}_{-ji} + w_j C^{[a]}_{j|-i} + \mathbb{1}_{a\geq 1}\cdot \alpha S^{[a-1]}_{-ji} - \big(\mathbb{1}_{a<A}\cdot\alpha + m^{[a]}\big) S^{[a]}_{-ji}$$

#### 4.3 Tertiary infection and fully immune states

- **Tertiary infection during cross-protection**

Transition: $I^{(C)[a]}_{k|-ji} \to R^{[a]}$

$$\frac{dI^{(C)[a]}_{k|-ji}}{dt} = \kappa_{jk}\eta_{ik}\lambda_k C^{[a]}_{j|-i} - \sigma I^{(C)[a]}_{k|-ji} + \mathbb{1}_{a\geq 1}\cdot \alpha I^{(C)[a-1]}_{k|-ji} - \big(\mathbb{1}_{a<A}\cdot\alpha + m^{[a]}\big) I^{(C)[a]}_{k|-ji}$$

- **Tertiary infection after waning of cross-protection**

Transition: $I^{(S)[a]}_{k|-ji} \to R^{[a]}$

$$\frac{dI^{(S)[a]}_{k|-ji}}{dt} = \max(\eta_{ik},\eta_{jk})\lambda_k S^{[a]}_{-ji} - \sigma I^{(S)[a]}_{k|-ji} + \mathbb{1}_{a\geq 1}\cdot \alpha I^{(S)[a-1]}_{k|-ji} - \big(\mathbb{1}_{a<A}\cdot\alpha + m^{[a]}\big) I^{(S)[a]}_{k|-ji}$$

- **Fully immune states**

Transition: $R^{[a]}$

$$\frac{dR^{[a]}}{dt} = \sum_{i}\sum_{j\neq i}\sum_{\substack{k\neq i\\k\neq j}} \sigma\big(I^{(C)[a]}_{k|-ji} + I^{(S)[a]}_{k|-ji}\big) + \mathbb{1}_{a\geq 1}\cdot \alpha R^{[a-1]} - \big(\mathbb{1}_{a<A}\cdot\alpha + m^{[a]}\big) R^{[a]}$$

Demographic processes follow the definitions in section 3, with births entering the youngest age group and ageing linking successive age classes. Indicator functions $\mathbb{1}_{a=0}$ and $\mathbb{1}_{a\geq 1}$ specify newborn inflow and ageing transitions, respectively. $A$ denotes the oldest age group (90 years and over). Birth and mortality rates are time-varying, and time arguments are omitted from the equations for clarity.

##### 4.4 Symptomatic cases

Symptomatic cases are calculated from new infections according to infection history, using the symptomatic probabilities in main text Table 1. For DENV, the symptomatic probability is $p_{D,pri}$ for primary infection and $p_{D,sec}$ for infection after prior DENV infection only. For DENV infection after prior ZIKV infection only, it is $\xi_{ZD}\, p_{D,pri}$. For DENV infection after prior infection with both DENV and ZIKV, the larger of $p_{D,sec}$ and $\xi_{ZD}\, p_{D,pri}$ applies.

For ZIKV, the symptomatic probability in age group $a$ at time $t$ is

$$p_Z^{[a]}(t) = p_{Z,g} + \big(p_{Z,cb} - p_{Z,g}\big)\, \theta^{[a]}(t),$$

where $\theta^{[a]}(t)$ is the proportion of pregnant women in age group $a$. ZIKV infection after prior DENV infection has symptomatic probability $\xi_{DZ}\, p_Z^{[a]}(t)$. The factors $\xi_{ZD}$ and $\xi_{DZ}$ are defined in main text Table 1 and section 2.2, and $\xi_{DZ}$ differs between the two symptomatic parameterizations.

The proportion $\theta^{[a]}(t)$ is approximated as $0.5 \times 0.75$ times the age-specific fertility rate for Thailand from the World Population Prospects [2] for ages 15–49 years, and is zero at other ages. The factor 0.5 approximates the female share of each age group, and 0.75 the duration of pregnancy as a share of a year.

### 5. Derivation of the reference basic reproduction number using the next-generation matrix approach

To characterize baseline transmission intensity, we derived a virus- or serotype-specific reference basic reproduction number ($R_0$) using the next-generation matrix (NGM) approach [1]. This derivation was carried out under a fully susceptible disease-free equilibrium (DFE), in which the population is infection-free and immunologically naive with respect to the invading virus or serotype. In this setting, all infections are primary infections. Individuals remaining susceptible to other virus or serotypes after prior infection are not included in this reference state, as their susceptibility may be modified by infection history and therefore does not correspond to full susceptibility. Secondary and subsequent infections, as well as cross-protection and enhancement, are therefore not involved in this derivation.

We focus on the primary-infection subsystem for a given virus or serotype $v$. Let $I_v^{[a]}$ denote infectious individuals in age group $a$, and define the total numbers of infectious individuals and susceptibles as

$$I_v = \sum_{a=0}^{A} I_v^{[a]}, \quad S = \sum_{a=0}^{A} S^{[a]}$$

The force of infection of virus or serotype $v$ is given by

$$\lambda_v = \frac{\rho_v \phi_v}{N} \sum_a I_v^{[a]} = \frac{\rho_v \phi_v}{N} I_v$$

Summing the equations for $I_v^{[a]}$ across age groups yields

$$\begin{aligned}
\frac{dI_v}{dt} &= \frac{d\left(\sum_{a=0}^{A} I_v^{[a]}\right)}{dt} = \sum_{a=0}^{A} \frac{dI_v^{[a]}}{dt} \\
&= \sum_{a=0}^{A} \left( \lambda_v S^{[a]} - \sigma I_v^{[a]} + \mathbb{1}_{a \geq 1} \cdot \alpha I_v^{[a-1]} - \left(\mathbb{1}_{a<A} \cdot \alpha + m^{[a]}\right) I_v^{[a]} \right) \\
&= \lambda_v S - \sigma I_v + \alpha \sum_{a=0}^{A-1} I_v^{[a]} - \alpha \sum_{a=0}^{A-1} I_v^{[a]} - \sum_{a=0}^{A} m^{[a]} I_v^{[a]} \\
&= \lambda_v S - \sigma I_v - \sum_{a=0}^{A} m^{[a]} I_v^{[a]}
\end{aligned}$$

During the early invasion phase, when the number of infectious individuals is small and the growth is approximately exponential, the age distribution of infectious individuals can be approximated by the age distribution of new infections. This approximation relies on homogeneous mixing and the age-independent form of the force of infection used in the model, under which the distribution of new infections across age groups is proportional to the susceptible population in each age group. It also relies on the infectious period being short relative to the time scales of ageing and mortality.

Under the fully susceptible DFE, all individuals are susceptible with respect to virus or serotype $v$, so that $S^{[a]} = N^{[a]}$. The age distribution of infections can therefore be approximated by the population age distribution:

$$\frac{I_v^{[a]}}{I_v} \approx \frac{N^{[a]}}{N}$$

Thus, under this approximation, the mortality term becomes

$$\begin{aligned}
\sum_{a=0}^{A} m^{[a]} I_v^{[a]} &= I_v \times \frac{1}{I_v} \sum_{a=0}^{A} m^{[a]} I_v^{[a]} \\
&= I_v \times \sum_{a=0}^{A} \frac{I_v^{[a]}}{I_v} m^{[a]} \\
&\approx I_v \times \sum_{a=0}^{A} \frac{N^{[a]}}{N} m^{[a]} \\
&= I_v \times \frac{\sum_{a=0}^{A} N^{[a]} m^{[a]}}{\sum_{a=0}^{A} N^{[a]}} \\
&:= I_v \times \bar{m}
\end{aligned}$$

where

$$\bar{m} = \frac{\sum_{a=0}^{A} N^{[a]} m^{[a]}}{\sum_{a=0}^{A} N^{[a]}}$$

denotes the age-weighted average mortality rate.

The age-weighted mortality rate was evaluated using age-specific mortality rates and population age structure at the start of the Thai demographic series, consistent with the demographic structure used at the start of the simulations.

Substituting this approximation into the aggregated equation gives

$$\frac{dI_v}{dt} \approx \lambda_v S - \big(\sigma + \bar{m}\big) I_v$$

At the DFE, $S = N$, and therefore

$$\frac{dI_v}{dt} = \lambda_v S - \big(\sigma + \bar{m}\big) I_v = \rho_v \phi_v I_v \frac{S}{N} - \big(\sigma + \bar{m}\big) I_v = \rho_v \phi_v I_v - \big(\sigma + \bar{m}\big) I_v$$

In the NGM framework, the infected state is one-dimensional, with state variable $x = I_v$, and we can rewrite its dynamics in the form:

$$\frac{dx}{dt} = \frac{dI_v}{dt} = F(I_v) - V(I_v)$$

where:

• $F(I_v)$ is the rate of new infections entering the aggregated infectious compartment $I_v$;

• $V(I_v)$ is the net rate of transfer out of $I_v$ due to all other processes including recovery and natural death. Ageing flows between age groups cancel in the age-aggregated equation and are therefore not treated as losses.

According to the linearized expression above, we identify

$$F(I_v) = \rho_v \phi_v I_v, \quad V(I_v) = \big(\sigma + \bar{m}\big) I_v$$

The corresponding Jacobians at the DFE are

$$F' = \frac{\partial F}{\partial I_v}\bigg|_{DFE} = \rho_v \phi_v, \quad V' = \frac{\partial V}{\partial I_v}\bigg|_{DFE} = \sigma + \bar{m}$$

Then the next-generation matrix $K$ reduces to a scalar:

$$K = \frac{F'}{V'} = \frac{\rho_v \phi_v}{\sigma + \bar{m}}$$

The basic reproduction number $R_{0,v}$ for virus or serotype $v$ is given by the spectral radius (dominant eigenvalue) of $K$. Since $K$ is a scalar, we obtain an approximation of $R_{0,v}$ in our system:

$$\boxed{R_{0,v} \approx \frac{\rho_v \phi_v}{\sigma + \bar{m}}}$$

This expression defines the reference basic reproduction number for virus or serotype $v$ under the fully susceptible DFE considered here. Note that this derivation reduces the full age-structured system to an age-aggregated one through the approximation that the age distribution of infectious individuals follows the population age distribution at the disease-free equilibrium. It is used to characterize baseline transmission intensity in the model, rather than to represent the reproduction threshold of the full cross-reactive transmission system. The age-structured dynamics emerge fully in the subsequent simulation of the ordinary differential equation system.

Given target values of $R_{0,v}$, baseline susceptibility parameter ($\rho_v$) was obtained by rearranging this expression:

$$\rho_v \approx \frac{R_{0,v}}{\phi_v} \times (\sigma + \bar{m})$$

Infectivity parameters ($\phi$) were set to 1 $d^{-1}$ for all virus or serotypes, such that baseline transmission intensity is fully captured by the susceptibility parameters ($\rho$). This is consistent with the model structure, in which cross-protection and enhancement are represented through modifications to susceptibility rather than infectivity. This normalization does not affect the resulting reproduction number, as transmission depends on the product of susceptibility and infectivity parameters.

#### 6. Initialization and warm-up of simulations

Each simulation was initialized from a 300-year warm-up under demographic conditions fixed at the first year of the Thai demographic series. The warm-up allowed the population immune structure to develop under sustained transmission before the main simulation began. Both DENV and ZIKV circulated during warm-up, and a separate warm-up was run for each cross-reactivity assumption. The state at the end of warm-up was used as the initial condition for the main simulation under time-varying demographic inputs.

For each cross-reactivity assumption, 100 simulations were run. One simulation used the values of $\rho_D$ and $\rho_Z$ in main text Table 1. In the other 99 simulations, $\rho_D$ and $\rho_Z$ were drawn from lognormal distributions, with $\log \rho_D \sim \mathcal{N}(-0.8478,\, 0.01^2)$ and $\log \rho_Z \sim \mathcal{N}(-1.5401,\, 0.01^2)$, whose medians are close to the values in main text Table 1. The same draws were used for all cross-reactivity assumptions. All other parameters were fixed at the values in main text Table 1.
