## Supplementary material 2: Supplementary figures and tables for "Impact of DENV-ZIKV cross-reactivity in Southeast Asia: A modelling study in Thailand"

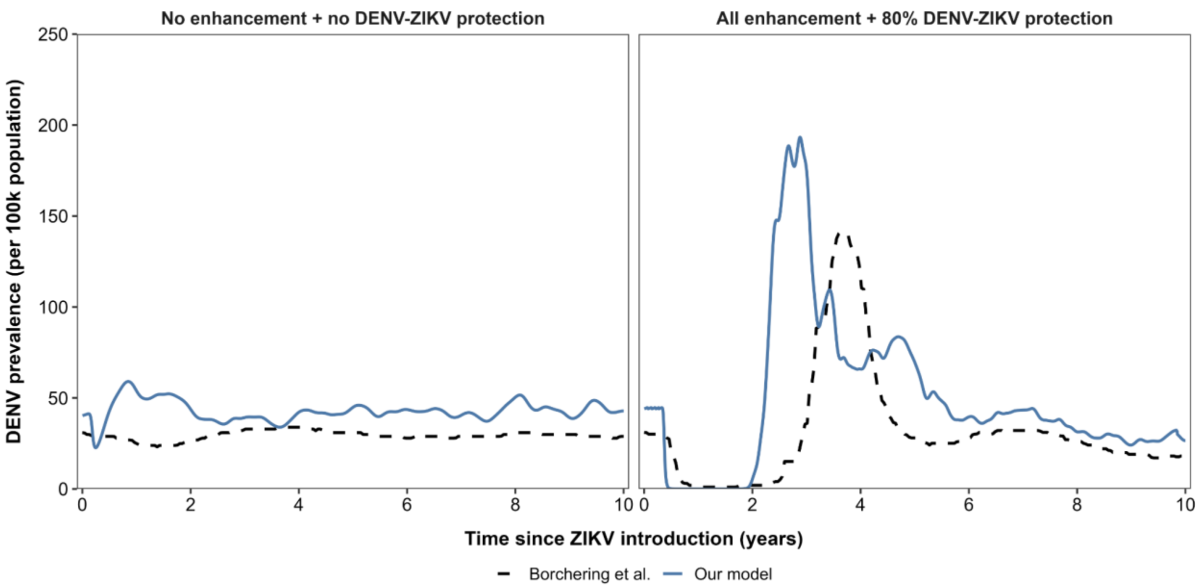


**Figure S1.** Benchmarking of post-ZIKV DENV dynamics against Borchering et al. Simulated mean DENV prevalence trajectories from the present model are compared with corresponding trajectories extracted from Borchering et al. under two cross-reactivity settings [1]. Solid lines show the mean trajectory across 100 simulations from the present model, and dashed lines show the corresponding mean trajectories from the reference study. The two settings are a setting without enhancement or DENV-ZIKV cross-protection (left panel) and an extreme cross-reactivity setting with enhancement in all directions and 80% DENV-ZIKV cross-protection (right panel). Borchering et al. used a stochastic model with explicit mosquito dynamics. The present model is deterministic and does not represent mosquito dynamics.

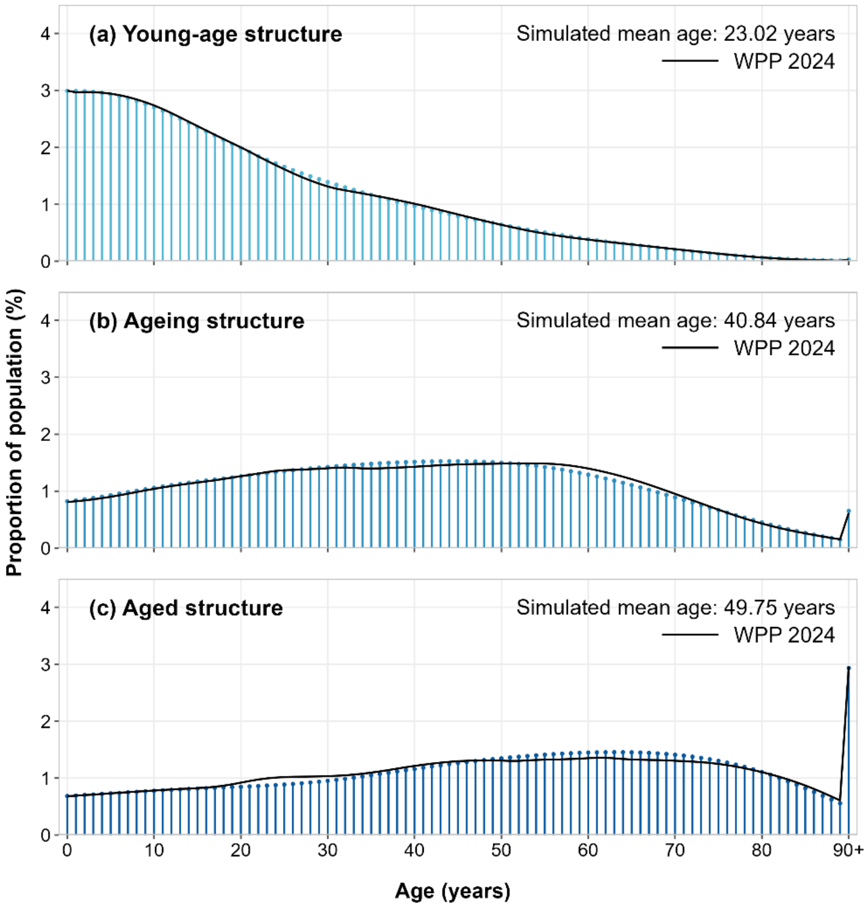


**Figure S2.** Age structure of the modelled Thai population. Needles show the proportion of the simulated population at each single year of age in the (a) young-age, (b) ageing and (c) aged demographic structures, averaged over the 10 years of each structure and based on Thai demographic parameters for the 1970s, 2020s and 2050s. Black lines show the corresponding age distributions from the World Population Prospects 2024 (WPP 2024) [2], using estimates for the 1970s and medium-variant projections for the 2020s and 2050s. The final age class includes individuals aged 90 years and over. The simulated mean age of the population is given in each panel. The corresponding mean ages in WPP 2024 are 23.05, 41.27 and 49.09 years. The simulated population closely reproduces the estimated and projected age structure of Thailand.

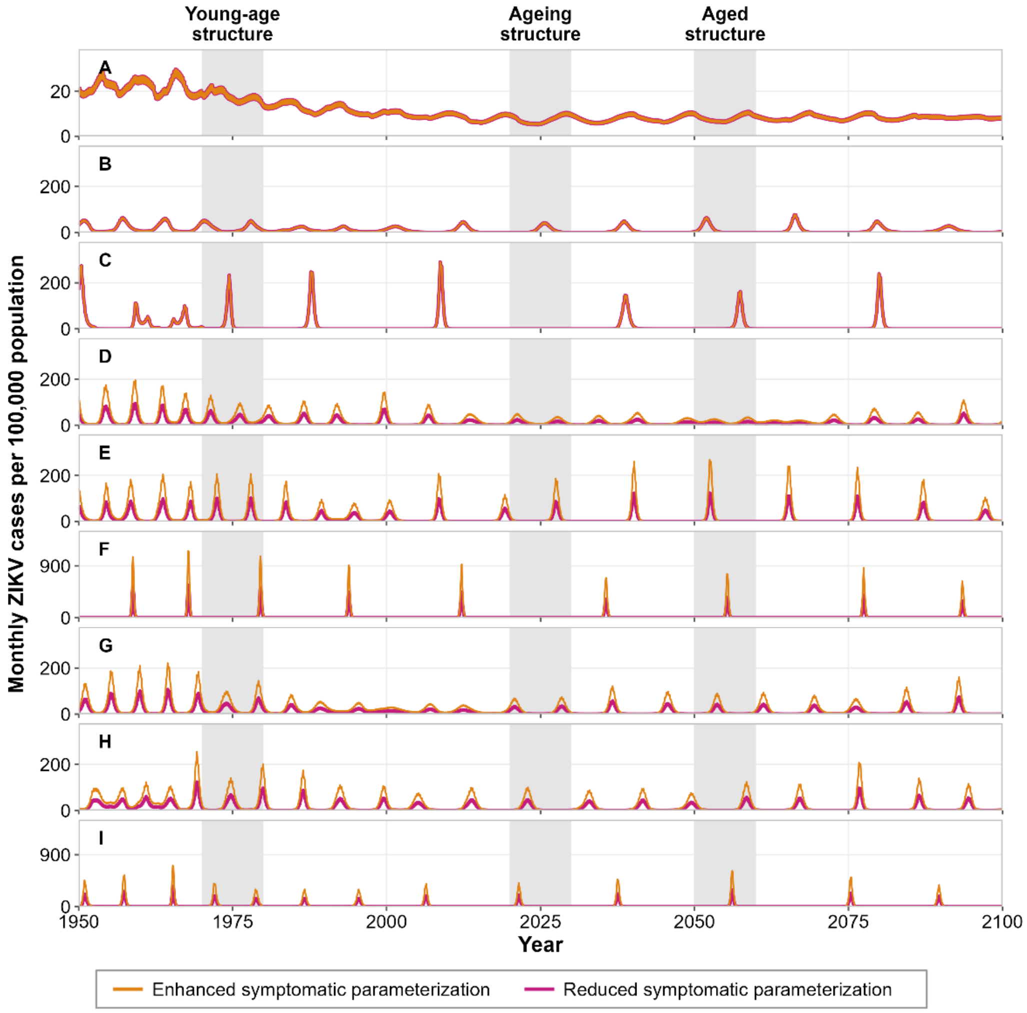


**Figure S3.** Monthly ZIKV cases in the two symptomatic parameterizations. Monthly ZIKV cases per 100,000 population from 1950 to 2099 under each cross-reactivity assumption, for the run with the parameter values in Table 1. Orange lines show the enhanced symptomatic parameterization and magenta lines the reduced symptomatic parameterization (section 2.2). Under assumptions A–C, the two parameterizations are identical and the lines coincide. Shaded bands mark the three 10-year windows used for the young-age, ageing and aged demographic structures. Y-axis ranges differ between assumption A, assumptions F and I, and the other assumptions. Rows are labelled by cross-reactivity assumption (Figure 1b).

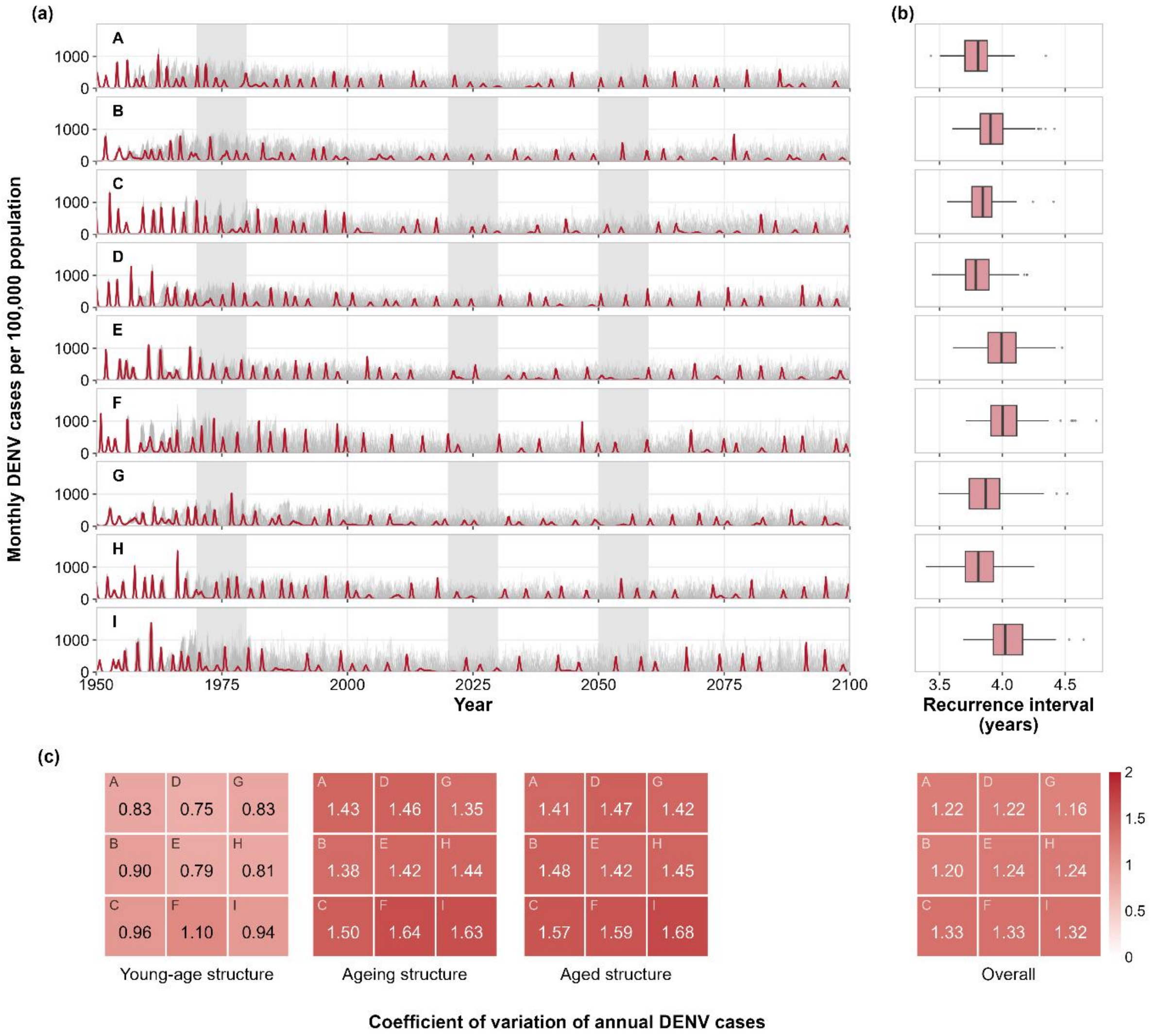


**Figure S4.** DENV transmission under sustained DENV-ZIKV co-circulation. (a) Monthly DENV cases per 100,000 population from 1950 to 2099 under each cross-reactivity assumption. Grey lines show individual runs and red lines the run with the parameter values in Table 1. Shaded bands mark the three 10-year windows used for the young-age, ageing and aged demographic structures. (b) Box plots of the recurrence interval of DENV epidemics across 100 simulation runs. (c) Heatmaps of the mean coefficient of variation of annual DENV cases across 100 simulation runs, within each demographic structure (below the corresponding windows in a) and over the full simulation period (Overall, below b). Panels in (a) and (b) and heatmap cells in (c) are labelled by cross-reactivity assumption (Figure 1b).

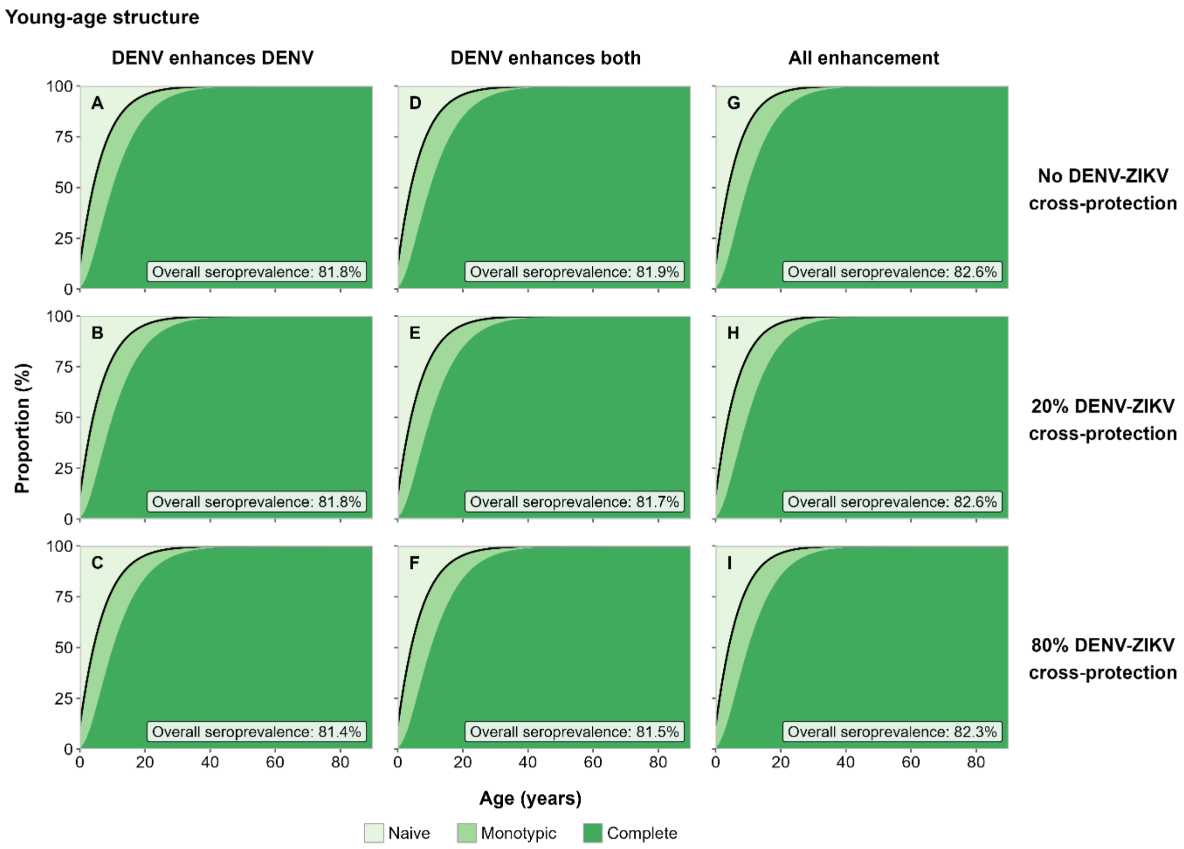


**Figure S5.** Age-specific DENV immunity under sustained co-circulation in the young-age structure. Proportions of individuals who are DENV-naive (light green), monotypically immune (medium green) or completely immune (dark green) by age, based on Thai demographic parameters for the 1970s. Columns show enhancement configurations and rows DENV-ZIKV cross-protection levels. The black line shows the proportion of individuals with at least one prior DENV infection, and each panel gives the overall DENV seroprevalence of the population. Proportions are means across 100 simulation runs. Panels are labelled by cross-reactivity assumption (Figure 1b).

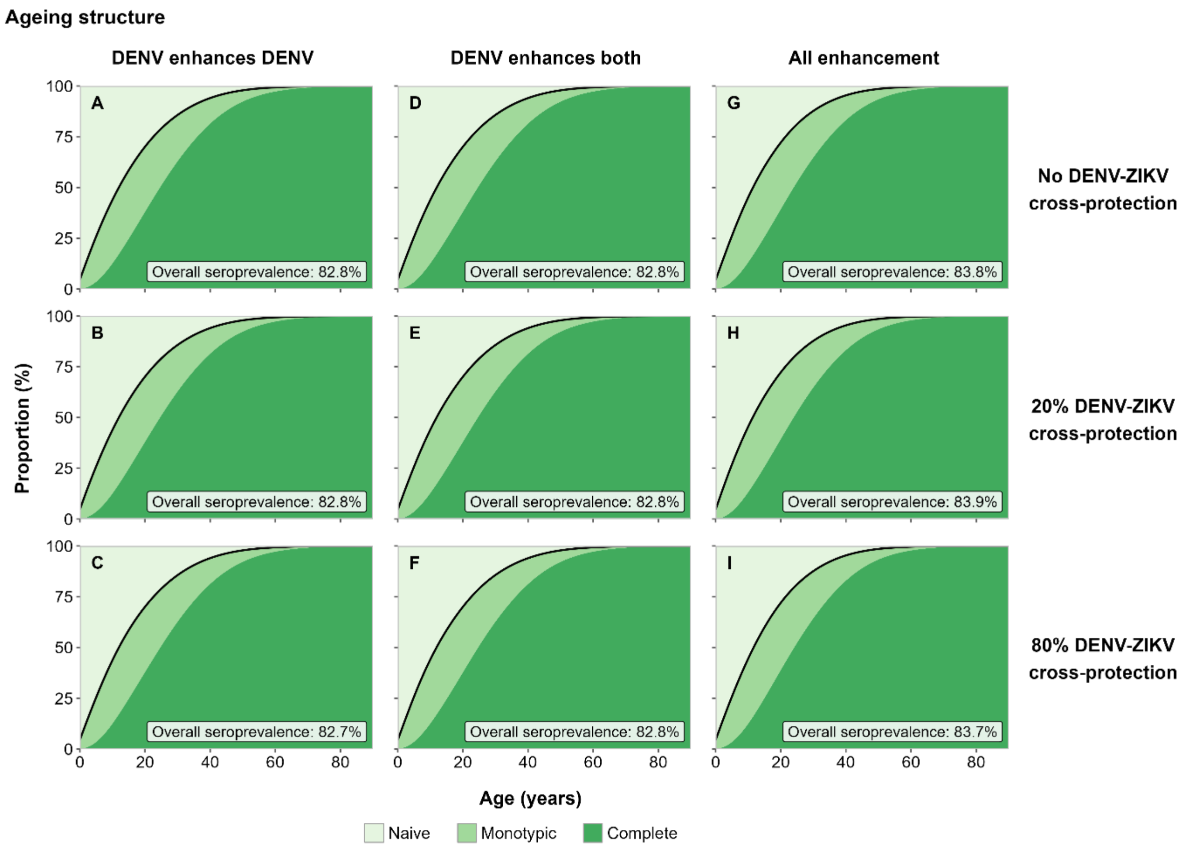


**Figure S6.** Age-specific DENV immunity under sustained co-circulation in the ageing structure. Proportions of individuals who are DENV-naive (light green), monotypically immune (medium green) or completely immune (dark green) by age, based on Thai demographic parameters for the 2020s. Columns show enhancement configurations and rows DENV-ZIKV cross-protection levels. The black line shows the proportion of individuals with at least one prior DENV infection, and each panel gives the overall DENV seroprevalence of the population. Proportions are means across 100 simulation runs. Panels are labelled by cross-reactivity assumption (Figure 1b).

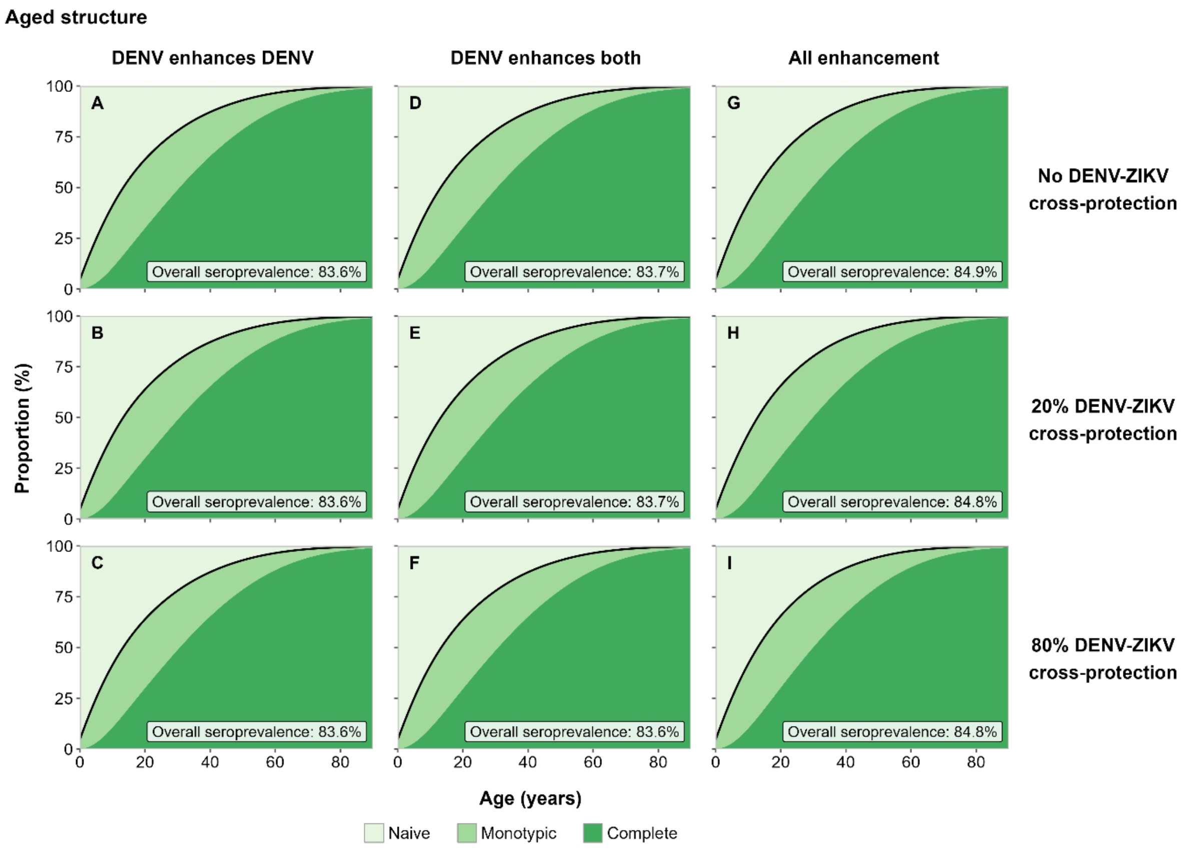


**Figure S7.** Age-specific DENV immunity under sustained co-circulation in the aged structure. Proportions of individuals who are DENV-naive (light green), monotypically immune (medium green) or completely immune (dark green) by age, based on Thai demographic parameters for the 2050s. Columns show enhancement configurations and rows DENV-ZIKV cross-protection levels. The black line shows the proportion of individuals with at least one prior DENV infection, and each panel gives the overall DENV seroprevalence of the population. Proportions are means across 100 simulation runs. Panels are labelled by cross-reactivity assumption (Figure 1b).

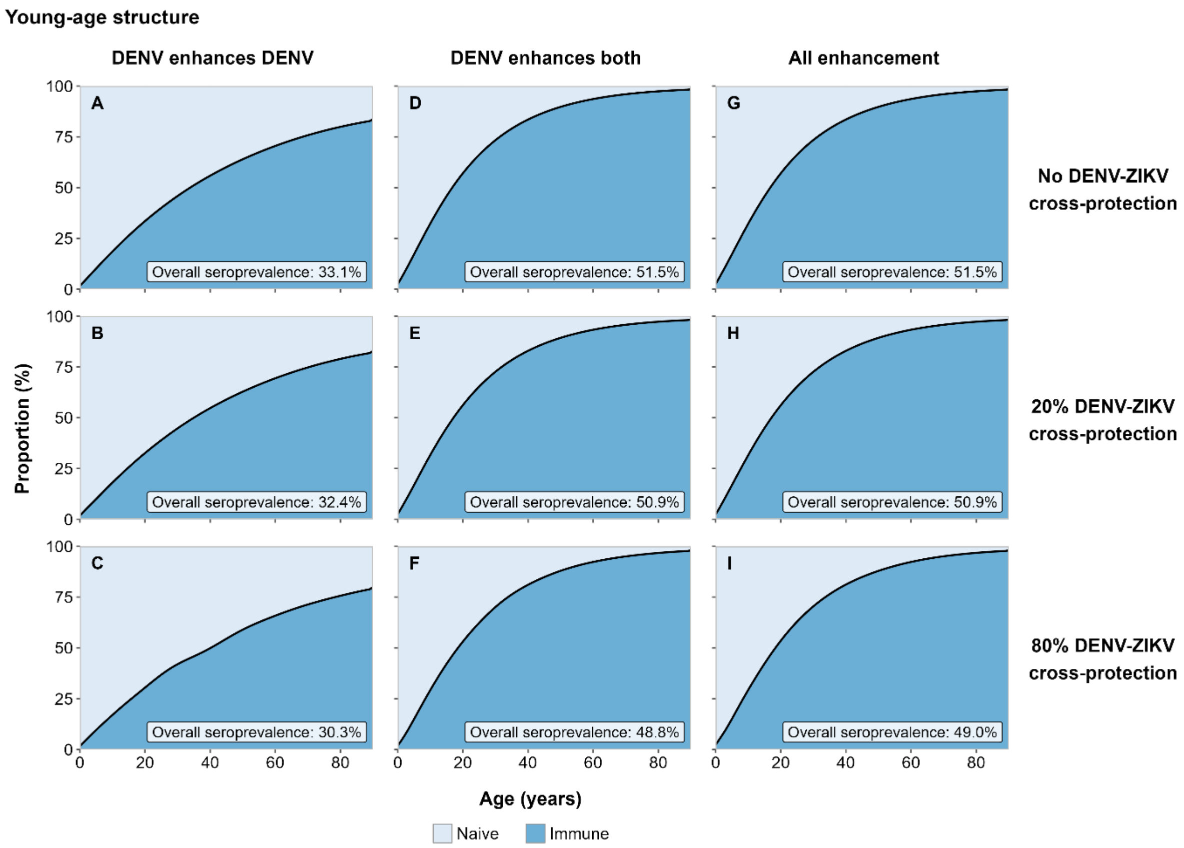


**Figure S8.** Age-specific ZIKV immunity under sustained co-circulation in the young-age structure. Proportions of individuals who are ZIKV-naive (light blue) or ZIKV-immune (dark blue) by age, based on Thai demographic parameters for the 1970s. Columns show enhancement configurations and rows DENV-ZIKV cross-protection levels. The black line shows the proportion of individuals with prior ZIKV infection, and each panel gives the overall ZIKV seroprevalence of the population. Proportions are means across 100 simulation runs. Panels are labelled by cross-reactivity assumption (Figure 1b).

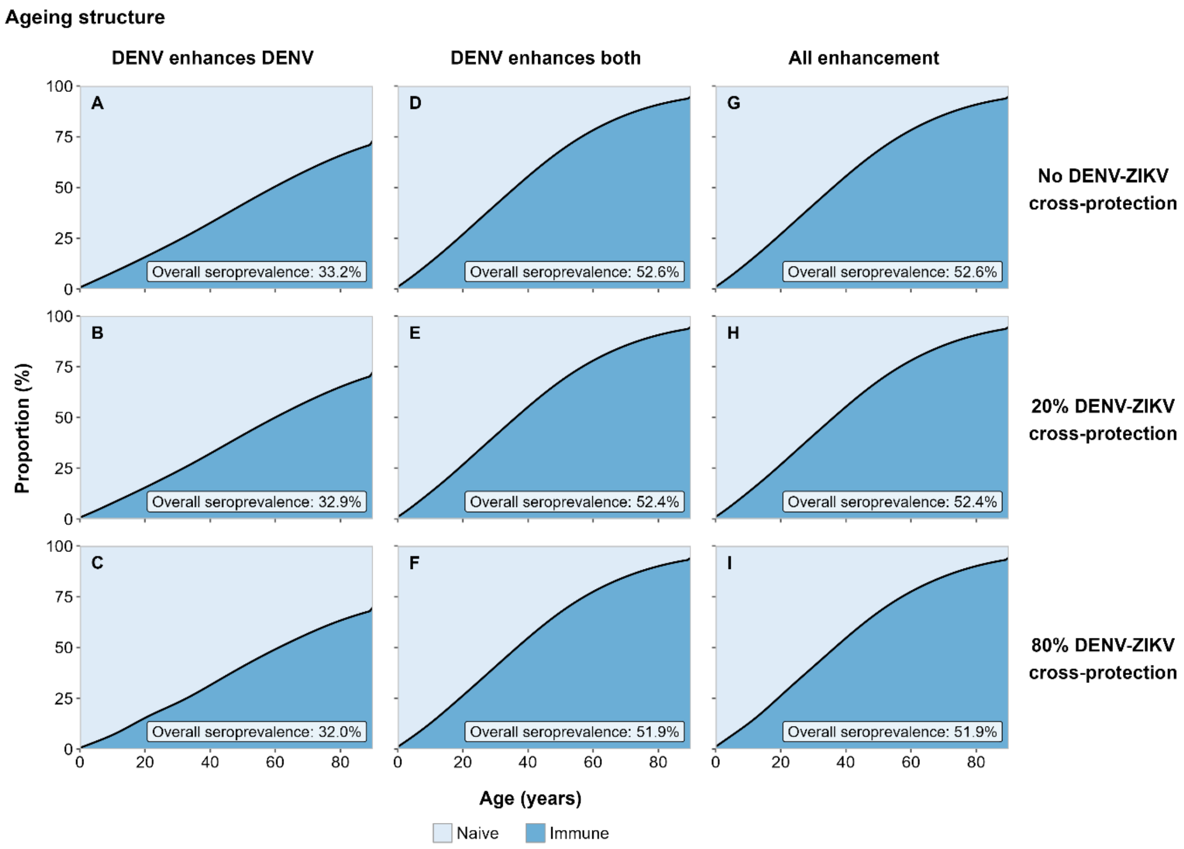


**Figure S9.** Age-specific ZIKV immunity under sustained co-circulation in the ageing structure. Proportions of individuals who are ZIKV-naive (light blue) or ZIKV-immune (dark blue) by age, based on Thai demographic parameters for the 2020s. Columns show enhancement configurations and rows DENV-ZIKV cross-protection levels. The black line shows the proportion of individuals with prior ZIKV infection, and each panel gives the overall ZIKV seroprevalence of the population. Proportions are means across 100 simulation runs. Panels are labelled by cross-reactivity assumption (Figure 1b).

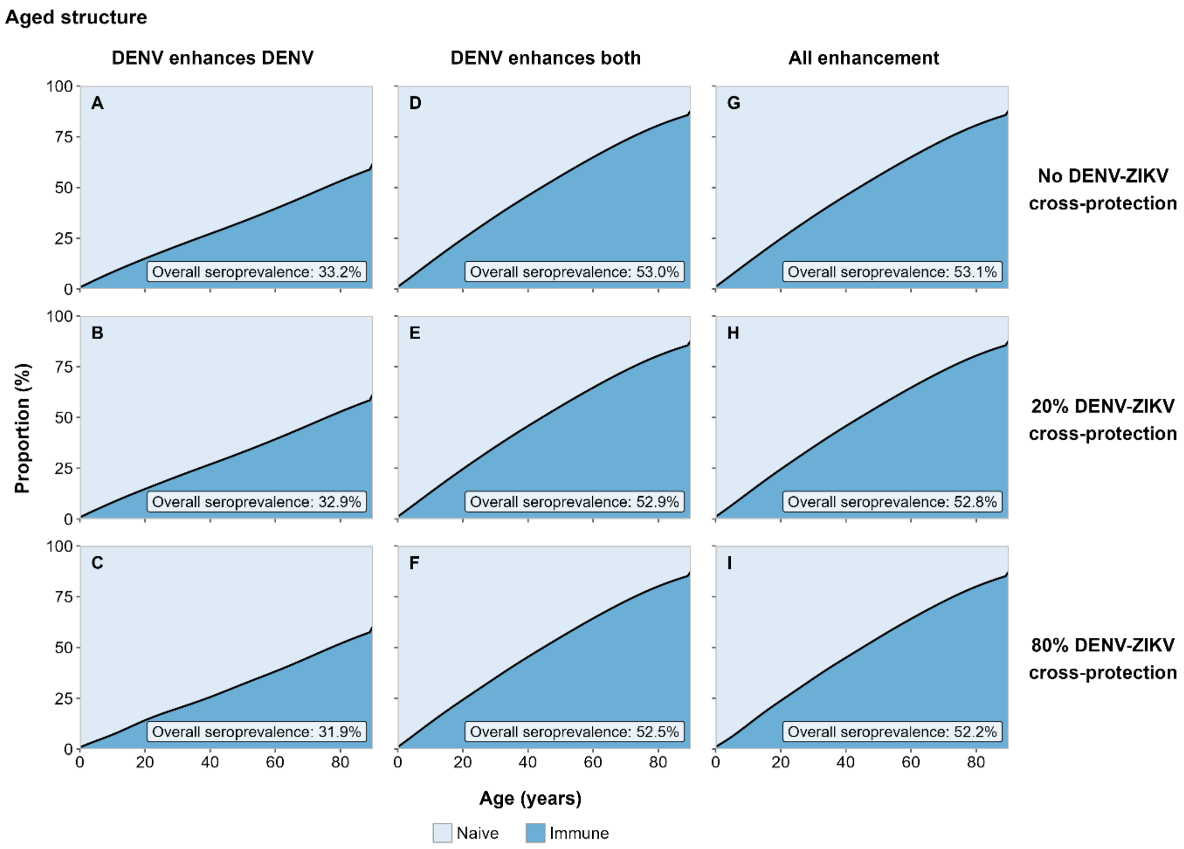


**Figure S10.** Age-specific ZIKV immunity under sustained co-circulation in the aged structure. Proportions of individuals who are ZIKV-naive (light blue) or ZIKV-immune (dark blue) by age, based on Thai demographic parameters for the 2050s. Columns show enhancement configurations and rows DENV-ZIKV cross-protection levels. The black line shows the proportion of individuals with prior ZIKV infection, and each panel gives the overall ZIKV seroprevalence of the population. Proportions are means across 100 simulation runs. Panels are labelled by cross-reactivity assumption (Figure 1b).

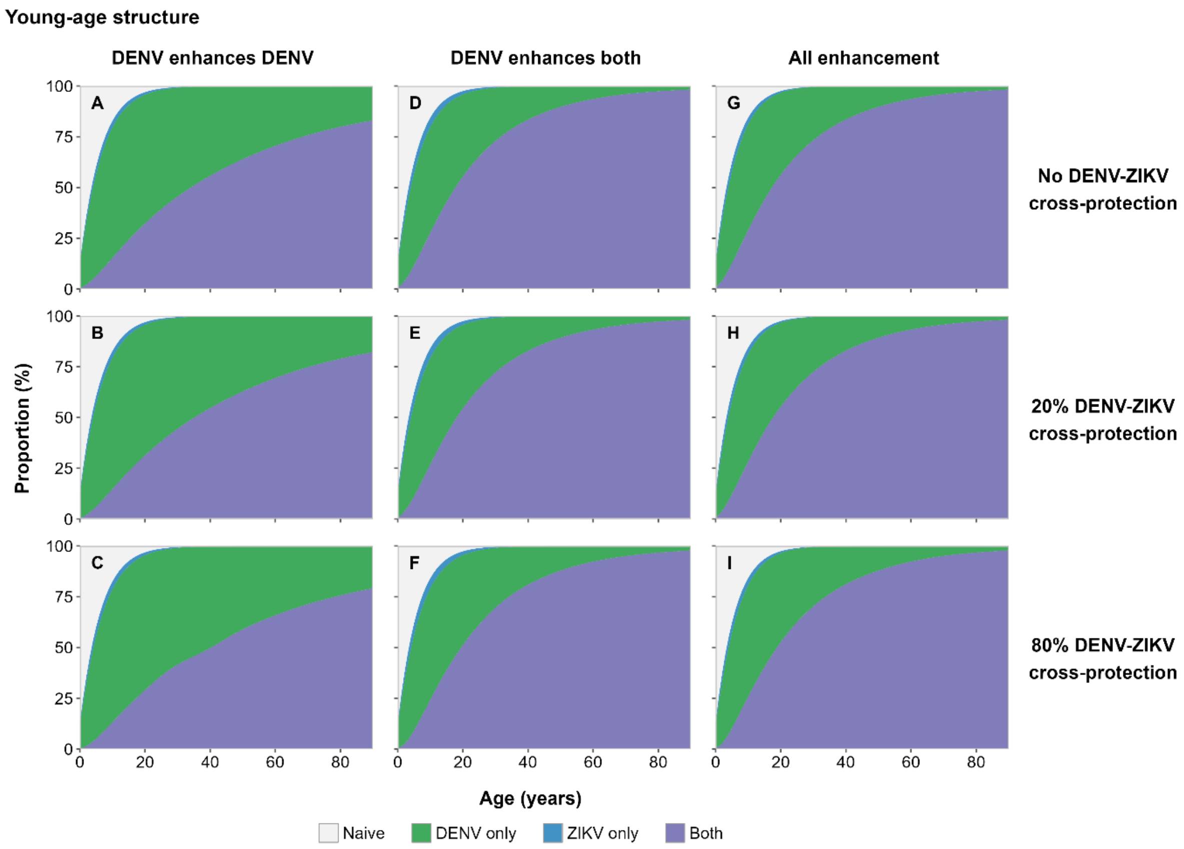


**Figure S11.** Age-specific immunity to DENV and ZIKV under sustained co-circulation in the young-age structure. Proportions of individuals who are naive to both viruses (grey), immune to DENV only (green), immune to ZIKV only (blue) or immune to both viruses (purple) by age, based on Thai demographic parameters for the 1970s. Columns show enhancement configurations and rows DENV-ZIKV cross-protection levels. Proportions are means across 100 simulation runs. Panel I is shown in Figure 3a. Panels are labelled by cross-reactivity assumption (Figure 1b).

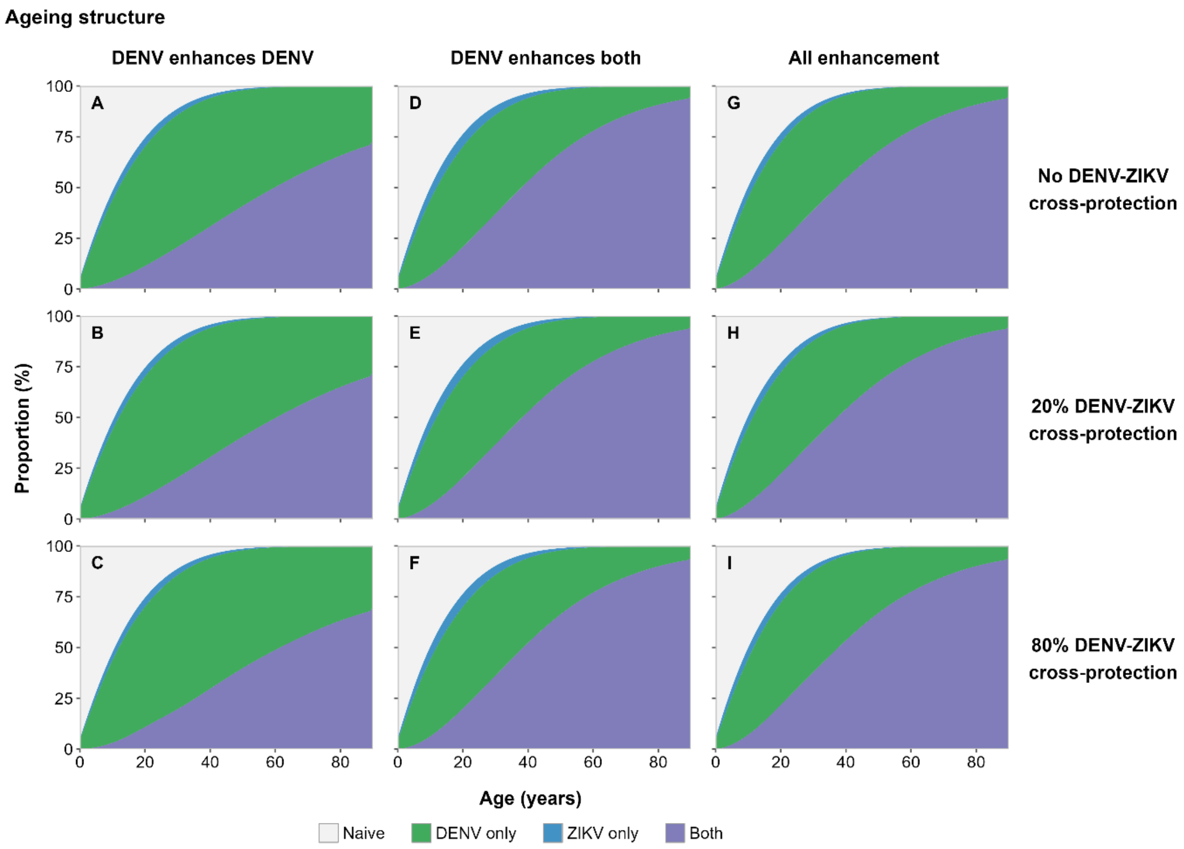


**Figure S12.** Age-specific immunity to DENV and ZIKV under sustained co-circulation in the ageing structure. Proportions of individuals who are naive to both viruses (grey), immune to DENV only (green), immune to ZIKV only (blue) or immune to both viruses (purple) by age, based on Thai demographic parameters for the 2020s. Columns show enhancement configurations and rows DENV-ZIKV cross-protection levels. Proportions are means across 100 simulation runs. Panel I is shown in Figure 3b. Panels are labelled by cross-reactivity assumption (Figure 1b).

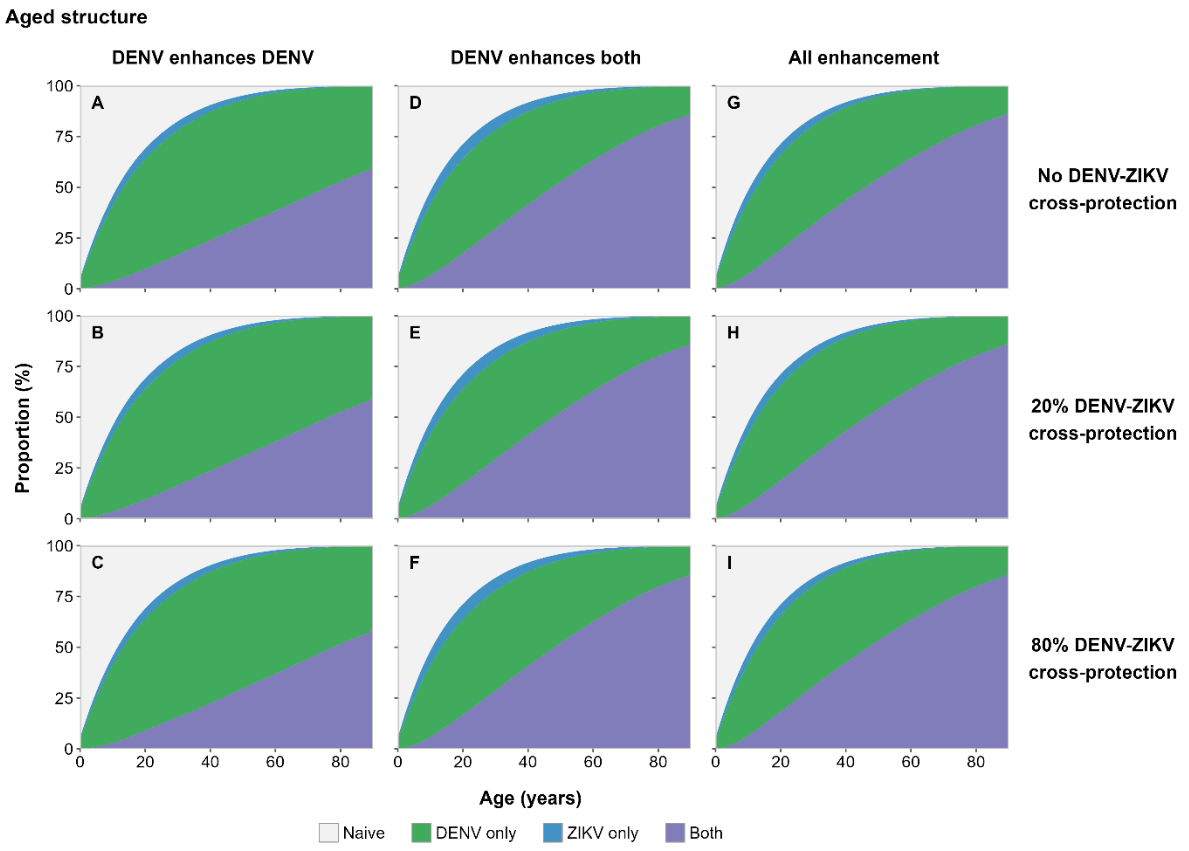


**Figure S13.** Age-specific immunity to DENV and ZIKV under sustained co-circulation in the aged structure. Proportions of individuals who are naive to both viruses (grey), immune to DENV only (green), immune to ZIKV only (blue) or immune to both viruses (purple) by age, based on Thai demographic parameters for the 2050s. Columns show enhancement configurations and rows DENV-ZIKV cross-protection levels. Proportions are means across 100 simulation runs. Panel I is shown in Figure 3c. Panels are labelled by cross-reactivity assumption (Figure 1b).

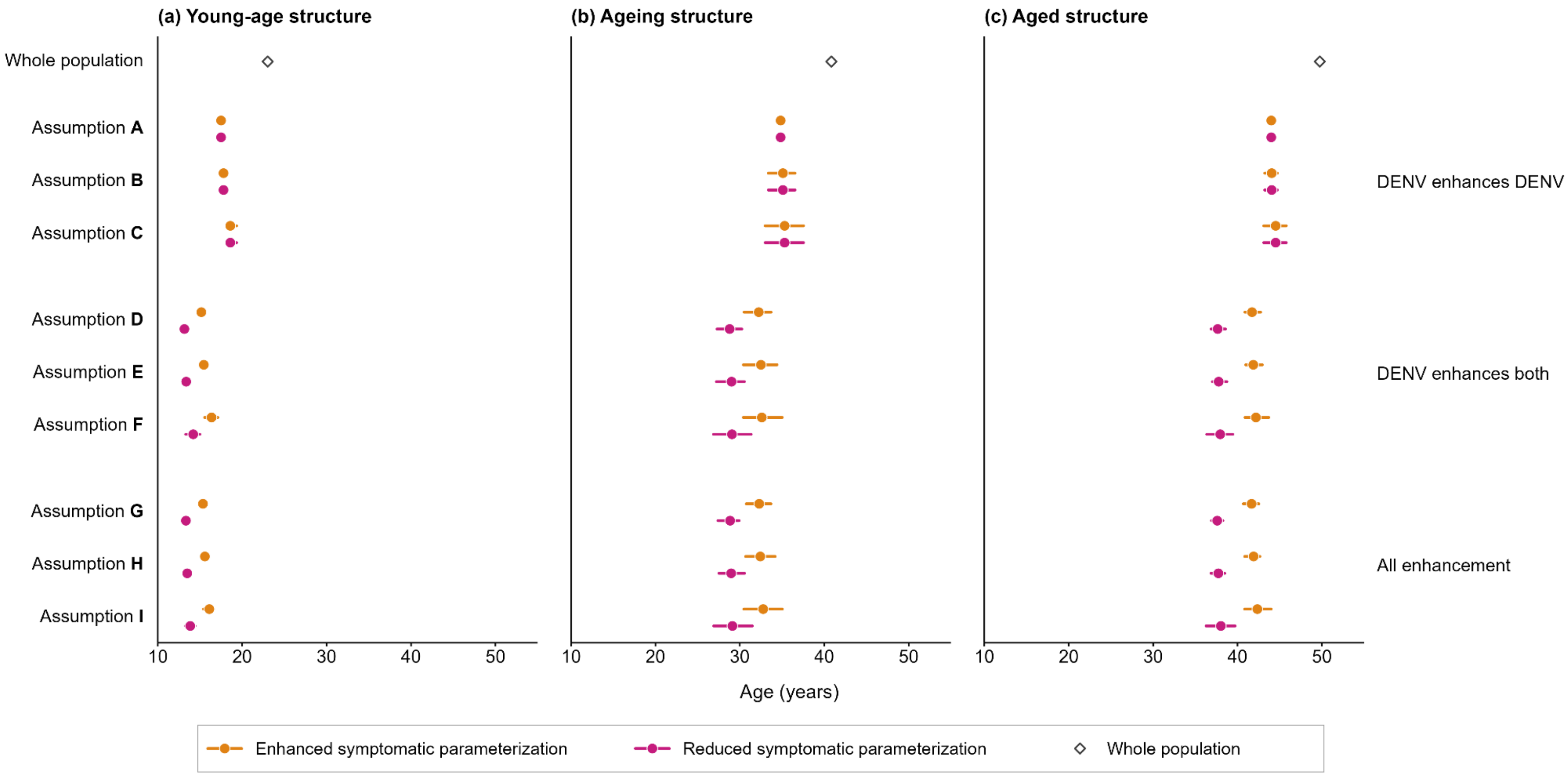


**Figure S14.** Mean age of ZIKV cases in the two symptomatic parameterizations. Points show the mean age of ZIKV cases across 100 runs under each cross-reactivity assumption, and horizontal lines show the 95% quantile range. Panels (a), (b) and (c) show the young-age, ageing and aged demographic structures, based on Thai demographic parameters for the 1970s, 2020s and 2050s. In each row, the upper point shows the enhanced symptomatic parameterization (orange) and the lower point the reduced symptomatic parameterization (magenta) (section 2.2). Under assumptions A–C, the two parameterizations are identical. Open diamonds show the mean age of the whole population in each structure. Assumptions are grouped by enhancement configuration (Figure 1b).

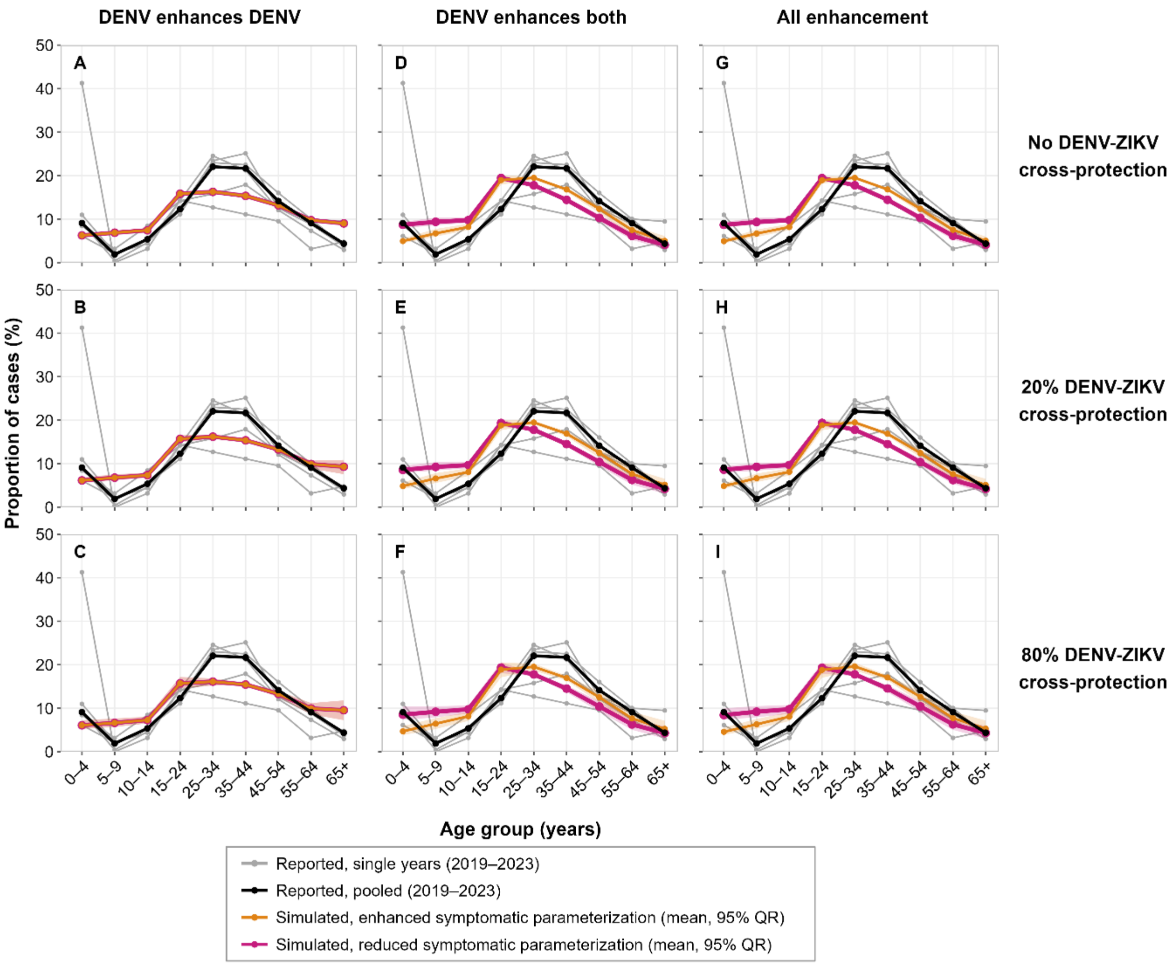


**Figure S15.** Simulated and reported age distributions of ZIKV cases in Thailand under all cross-reactivity assumptions. Grey lines show the reported age distributions in each year from 2019 to 2023, and the black line the distribution pooled across the five years. Orange and magenta lines show the mean age distributions of simulated ZIKV cases across 100 runs in the ageing structure, in the enhanced and reduced symptomatic parameterizations (section 2.2). Shading shows the 95% quantile range across runs. Under assumptions A–C, the two parameterizations are identical and the lines coincide. Panels are labelled by cross-reactivity assumption (Figure 1b).

**Table S1.** Average annual DENV and ZIKV cases under sustained co-circulation. Average annual number of DENV and ZIKV cases per 100,000 population from 1950 to 2099 under each cross-reactivity assumption. ZIKV cases are given for the enhanced and reduced symptomatic parameterizations (section 2.2). DENV cases are the same in both parameterizations. Values are means across 100 simulation runs, with the 95% quantile range (QR). Rows are labelled by cross-reactivity assumption (Figure 1b).

| Enhancement configuration | DENV-ZIKV cross-protection | Assumption | Mean annual cases per 100k population | | |
|---|---|---|---|---|---|
| | | | DENV | ZIKV, by symptomatic parameterization | |
| | | | | Enhanced | Reduced |
| | | | mean (95% QR) | mean (95% QR) | mean (95% QR) |
| DENV enhances DENV | None | A | 794.79 (784.13, 805.14) | 131.51 (127.23, 137.23) | 131.51 (127.23, 137.23) |
| | 20% | B | 791.07 (782.16, 801.10) | 131.43 (125.13, 138.98) | 131.43 (125.13, 138.98) |
| | 80% | C | 792.03 (781.86, 804.47) | 138.07 (128.25, 150.11) | 138.07 (128.25, 150.11) |
| DENV enhances both | None | D | 792.82 (780.02, 803.04) | 284.75 (278.29, 291.71) | 136.32 (133.17, 139.45) |
| | 20% | E | 782.48 (773.39, 790.88) | 285.03 (277.44, 294.83) | 136.68 (133.26, 140.71) |
| | 80% | F | 804.34 (789.15, 815.77) | 270.01 (257.35, 286.46) | 129.44 (123.24, 136.93) |
| All enhancement | None | G | 814.24 (801.46, 823.64) | 290.89 (285.15, 297.21) | 139.08 (136.53, 142.21) |
| | 20% | H | 822.07 (812.74, 831.20) | 284.68 (276.92, 293.12) | 136.55 (132.93, 140.31) |
| | 80% | I | 816.86 (800.32, 828.72) | 292.99 (279.03, 307.88) | 141.03 (134.74, 147.85) |

**Table S2. Mean age of DENV and ZIKV cases under sustained co-circulation.** Mean age (years) of DENV and ZIKV cases under each cross-reactivity assumption in the young-age, ageing and aged demographic structures, based on Thai demographic parameters for the 1970s, 2020s and 2050s. Values are means across 100 simulation runs, with the 95% quantile range (QR). The mean age of the whole population in each structure is given for reference. ZIKV values are for the enhanced symptomatic parameterization (section 2.2). Rows are labelled by cross-reactivity assumption (Figure 1b).

| Enhancement configuration | DENV-ZIKV cross-protection | Assumption | Mean age of cases (years) | | | | | |
|---|---|---|---|---|---|---|---|---|
| | | | Young-age structure | | Ageing structure | | Aged structure | |
| | | | DENV | ZIKV | DENV | ZIKV | DENV | ZIKV |
| | | | mean (95% QR) | mean (95% QR) | mean (95% QR) | mean (95% QR) | mean (95% QR) | mean (95% QR) |
| Whole population (reference) | – | – | 23.02 | | 40.84 | | 49.75 | |
| DENV enhances DENV | None | A | 9.17 (8.97, 9.35) | 17.50 (17.26, 17.66) | 22.56 (21.58, 23.95) | 34.83 (34.55, 35.31) | 30.51 (29.53, 31.42) | 44.00 (43.68, 44.34) |
| | 20% | B | 9.20 (9.03, 9.36) | 17.79 (17.48, 17.95) | 22.60 (21.60, 23.82) | 35.10 (33.37, 36.54) | 30.52 (29.53, 31.53) | 44.05 (43.22, 44.73) |
| | 80% | C | 9.27 (8.87, 9.54) | 18.59 (18.12, 19.34) | 22.62 (21.28, 23.73) | 35.30 (33.00, 37.55) | 30.53 (29.49, 31.59) | 44.52 (43.11, 45.78) |
| DENV enhances both | None | D | 9.16 (8.97, 9.32) | 15.16 (14.90, 15.46) | 22.61 (21.59, 23.61) | 32.24 (30.50, 33.70) | 30.42 (29.52, 31.30) | 41.72 (40.87, 42.74) |
| | 20% | E | 9.23 (8.88, 9.43) | 15.45 (15.12, 15.69) | 22.62 (21.58, 23.96) | 32.50 (30.42, 34.40) | 30.46 (29.49, 31.31) | 41.87 (40.97, 42.94) |
| | 80% | F | 9.32 (9.05, 9.60) | 16.37 (15.55, 17.13) | 22.66 (21.24, 24.85) | 32.60 (30.40, 35.03) | 30.72 (29.79, 32.43) | 42.18 (40.89, 43.71) |
| All enhancement | None | G | 9.01 (8.73, 9.14) | 15.35 (15.10, 15.51) | 22.10 (21.23, 23.05) | 32.29 (30.75, 33.68) | 29.68 (28.66, 30.36) | 41.67 (40.68, 42.53) |
| | 20% | H | 9.01 (8.78, 9.23) | 15.58 (15.39, 15.71) | 22.04 (20.91, 23.19) | 32.42 (30.68, 34.19) | 29.82 (29.07, 30.90) | 41.90 (40.84, 42.65) |
| | 80% | I | 9.15 (8.91, 9.43) | 16.10 (15.41, 16.61) | 22.31 (20.90, 24.51) | 32.76 (30.47, 35.04) | 29.95 (29.01, 30.92) | 42.35 (40.84, 43.99) |

**Table S3. Distance between simulated and reported age distributions of ZIKV cases in Thailand.** Total variation distance (TVD) and root mean squared error (RMSE, %) between the age distribution of simulated ZIKV cases in the ageing structure and the reported Thai distribution pooled across 2019–2023, using nine age groups, under each cross-reactivity assumption. Distances are given for the enhanced and reduced symptomatic parameterizations (section 2.2). Values are means across 100 simulation runs, with the 95% quantile range (QR). Rows are labelled by cross-reactivity assumption (Figure 1b).

| Enhancement configuration | DENV-ZIKV cross-protection | Assumption | Distance from the reported age distribution of ZIKV cases | | | |
|---|---|---|---|---|---|---|
| | | | Enhanced symptomatic parameterization | | Reduced symptomatic parameterization | |
| | | | TVD mean (95% QR) | RMSE (%) mean (95% QR) | TVD mean (95% QR) | RMSE (%) mean (95% QR) |
| DENV enhances DENV | None | A | 0.159 (0.158, 0.161) | 4.03 (4.01, 4.07) | 0.159 (0.158, 0.161) | 4.03 (4.01, 4.07) |
| | 20% | B | 0.160 (0.156, 0.166) | 4.07 (3.98, 4.27) | 0.160 (0.156, 0.166) | 4.07 (3.98, 4.27) |
| | 80% | C | 0.161 (0.150, 0.172) | 4.15 (3.83, 4.50) | 0.161 (0.150, 0.172) | 4.15 (3.83, 4.50) |
| DENV enhances both | None | D | 0.150 (0.137, 0.171) | 3.80 (3.48, 4.30) | 0.191 (0.176, 0.217) | 4.96 (4.49, 5.51) |
| | 20% | E | 0.147 (0.129, 0.173) | 3.74 (3.34, 4.36) | 0.189 (0.173, 0.218) | 4.88 (4.41, 5.54) |
| | 80% | F | 0.149 (0.129, 0.173) | 3.80 (3.33, 4.36) | 0.193 (0.168, 0.225) | 4.91 (4.21, 5.64) |
| All enhancement | None | G | 0.149 (0.136, 0.168) | 3.78 (3.45, 4.25) | 0.190 (0.178, 0.215) | 4.94 (4.59, 5.47) |
| | 20% | H | 0.148 (0.132, 0.170) | 3.76 (3.39, 4.29) | 0.189 (0.172, 0.212) | 4.90 (4.39, 5.42) |
| | 80% | I | 0.146 (0.128, 0.171) | 3.74 (3.33, 4.32) | 0.192 (0.165, 0.226) | 4.89 (4.13, 5.69) |

**Table S4. Reported ZIKV cases in Thailand by age group, 2019–2023.** Number of reported ZIKV cases in each reported age group and year, the total over 2019–2023 and the pooled proportion (%) of cases in each age group. Data are from the Bureau of Epidemiology, Department of Disease Control, Ministry of Public Health, Thailand [3]. "<1" denotes infants under 1 year of age.

| Age group (years) | Reported ZIKV cases | | | | | | Pooled proportion (%) |
|---|---|---|---|---|---|---|---|
| | 2019 | 2020 | 2021 | 2022 | 2023 | Total | |
| <1 | 26 | 18 | 25 | 7 | 34 | 110 | 6.9 |
| 1 | 1 | 0 | 0 | 2 | 4 | 7 | 0.4 |
| 2 | 1 | 1 | 1 | 0 | 5 | 8 | 0.5 |
| 3 | 1 | 1 | 0 | 4 | 5 | 11 | 0.7 |
| 4 | 1 | 2 | 0 | 3 | 2 | 8 | 0.5 |
| 5 | 3 | 0 | 0 | 0 | 2 | 5 | 0.3 |
| 6 | 0 | 0 | 0 | 1 | 5 | 6 | 0.4 |
| 7–9 | 3 | 1 | 0 | 5 | 10 | 19 | 1.2 |
| 10–14 | 12 | 11 | 2 | 16 | 44 | 85 | 5.4 |
| 15–24 | 39 | 29 | 9 | 27 | 91 | 195 | 12.3 |
| 25–34 | 67 | 56 | 8 | 30 | 188 | 349 | 22.0 |
| 35–44 | 58 | 60 | 7 | 34 | 184 | 343 | 21.7 |
| 45–54 | 33 | 30 | 6 | 24 | 131 | 224 | 14.2 |
| 55–64 | 20 | 21 | 2 | 19 | 82 | 144 | 9.1 |
| 65+ | 8 | 9 | 3 | 18 | 31 | 69 | 4.4 |
| Total | 273 | 239 | 63 | 190 | 818 | 1583 | 100.0 |

[2] United Nations, Department of Economic and Social Affairs, Population Division. 2024 World Population Prospects 2024, Data Sources.

[3] Bureau of Epidemiology, Department of Disease Control, Ministry of Public Health, Thailand. 2025 Zika: Annual situation summary. Dep. Dis. Control MoPH Thail. See http://doe1.moph.go.th/surdata/disease.php?dcontent=old&ds=87 (accessed on 20 August 2025).
